# Chronic heart failure associates with enhanced activation and clonal expansion of T cells with predicted autoreactive capacity

**DOI:** 10.1101/2025.04.14.25325772

**Authors:** Maximilian Merten, Tina Rasper, Kathrin Holz, Sebastian Cremer, Bianca Schuhmacher, Lukas Tombor, David John, Evelyn Ullrich, Igor Macinkovic, David Leistner, Jedrzej Hoffmann, Timotheus Speer, Halvard Bonig, Christoph Knosalla, Evgenij Potapov, Tamim Sarapki, Emina Mustafic, Alexander Berkowitsch, Andreas Michael Zeiher, Stefanie Dimmeler, Wesley Tyler Abplanalp

## Abstract

Chronic heart failure (HF) is characterized by adverse remodeling and persistent inflammation, contributing to impaired heart function and poor prognosis. While the acute immune response post-myocardial infarction (MI) is well-studied, its role in chronic HF remains unclear.

Phenotyping of peripheral blood T cells by flow cytometry and single-cell RNA sequencing revealed T central memory cells (TCM) decline, while CD4^+^ Th17 and CD8^+^ T effector memory cells increase in HF patients compared to healthy age-matched controls. Furthermore, this decline in TCM cells and increase in homing marker CCR5 on T cell subsets associates with poor prognosis in HF. T cell receptor sequencing revealed clonal expansion in circulating and cardiac T cells, while epitope prediction modeling suggested autoreactivity of T cells in HF. Spatial and scRNA-seq data confirm inflammatory T cell infiltration in the human and murine heart post-MI.

In summary, HF shows potential autoreactivity, with an increased homing capacity and declining TCM cells associated with poor prognosis.

## Introduction

Chronic ischemic heart failure (HF) represents a global health challenge characterized by adverse cardiac remodeling and persistent inflammation which can lead to impaired left ventricular (LV) function. Despite initial indications from pre-clinical and early human studies proposing anti-inflammatory therapies for treatment of HF, large-scale clinical trials involving antibody-based cytokine neutralization of TNFα failed to demonstrate significant benefits.^1^ Promisingly, targeting IL1β with canakinumab (CANTOS Trial) demonstrated modest reduction in cardiovascular risk in a group of highly inflamed patients.^2,3^ As immunomodulation therapies targeting HF can present increased risk for opportunistic infection, there is a need for more precise identification of inflammatory targets in HF patients. Rather than relying solely on plasma cytokine levels, in depth profiling of immune subsets may provide more insights into the specific alterations of immune cell populations and their contributions to heart failure.^4^

Recent findings in chronic ischemic HF revealed a robust expansion of monocytes/macrophages and dendritic cells, forming a pro-inflammatory phenotype. These observations prompt consideration for directly targeting specific leukocyte populations as an effective alternative strategy for immunomodulation in HF. Dendritic cells and to some extent macrophages play a pivotal role in antigen presentation to T cells, thereby initiating T cell activation.^5^ The documented global expansion of dendritic cells and monocytes/macrophages in HF suggests a parallel activation of T cells. Various T cell populations, such as CD4^+^ T cells, Foxp3^+^ regulatory T cells (Tregs), and invariant natural killer T cells have demonstrated favorable roles in inflammation regulation, immune cell trafficking, angiogenesis, and cardiac remodeling in the context of acute myocardial infarction (MI).^6^ However, the impact of T cell modulation in chronic ischemic HF remains unclear.

In murine models of non-ischemic pressure-overload induced HF, activated CD4^+^ T-cells have been associated with cardiac fibrosis, hypertrophy, and adverse remodeling.^7^ Altered circulating lymphocyte counts have also been shown to predict worse outcomes in HF patients. ^8^ However, the specific impact of lymphocyte subpopulations, particularly in relation to HF disease stage, remains poorly understood.

CD4^+^ and CD8^+^ T cells comprise a diverse family of adaptive immune cells with distinct functions, including pro-inflammatory T helper (Th)1 cells, anti-inflammatory Th2 cells, pro-inflammatory Th17 cells, and immunomodulatory Tregs.^9^ Given the chronic inflammatory nature of ischemic HF, alterations in the activation and distribution of these T cell subsets may significantly influence the progression of pathological LV remodeling.^10^ Recent studies in patients and mice with pronounced atherosclerosis show a shift of Tregs towards the more pro-inflammatory status in the peripheral blood, displaying increased CD16 and CD56 cytotoxic markers.^11^ Nevertheless, it is widely unclear how these immune cells interact and change during the course of the disease.

Here we addressed these open questions using flow cytometry and single-cell RNA sequencing (scRNA-seq) of peripheral blood cells derived from HF patients and age matched healthy controls. We show pronounced T cell activation, clonal expansion, and human self-targeting epitopes in HF patients, along with prediction for mortality. Additionally, our research elucidates long term temporal dynamics of post-MI T cell responses in a murine model, offering insights into the pro-inflammatory T cell landscape during different phases of HF progression.

## Results

### Chronic HF associates with latent T cell activation and clonal expansion

To study immunological consequences of HF, we analyzed T cells from whole blood of 44 healthy controls (HC) and 144 HF patients by flow cytometry (Extended Data Tables 1 and 2). Controls were matched for age and had a mixed sex background. Average age was 65.0 years for HC and 67.5 years for HF (p=0.09). Proportion of male donors were 50.0% and 85.4% in the HC and HF populations, respectively. Within the HF population, LVEF (left ventricular ejection fraction) was 28.2 ± 9.6%, GFR (Glomerular filtration rate) was 53.7 ± 21.7, and CRP (C-reactive protein) was 0.8 ± 1.6 mg/dL, with mean follow up time of 453.8 ± 242.0 days. Serum analysis by Legendplex assay of HF and HC derived samples showed significantly elevated levels of IL6, sTNF-RI and sICAM-1 in HF patients, indicating the presence of inflammation and tissue damage (Extended Data Figure 1a).

HF patients had a significant increase in combined memory/effector and TEM (T effector memory) CD8^+^ T cell frequencies relative to HC (Figure 1a), whereas CD4^+^ and CD8^+^ T central memory cells (TCM) were reduced in HF (Figure 1a,b). Frequencies of a CD4^+^ or CD8^+^ subset, always refer to total CD4^+^ or CD8^+^ T cell parent populations, respectively. T effector memory cells are direct mediators of an immune response and have higher tissue specificity and capacity to enter tissue sites. Analysis of CD4^+^ T helper subsets revealed significant phenotypical shifts with a relative decrease of Th1 cells in HF patients, while Th17 and Treg cells were increased in HF compared to HC (Figure 1c). Interestingly, subanalysis of Treg cells (Extended Data 1b-g) showed increased Th17-like and Th9-like Treg skewing, suggesting HF Tregs may have diminished immunosuppressive activity.

**Figure 1:**
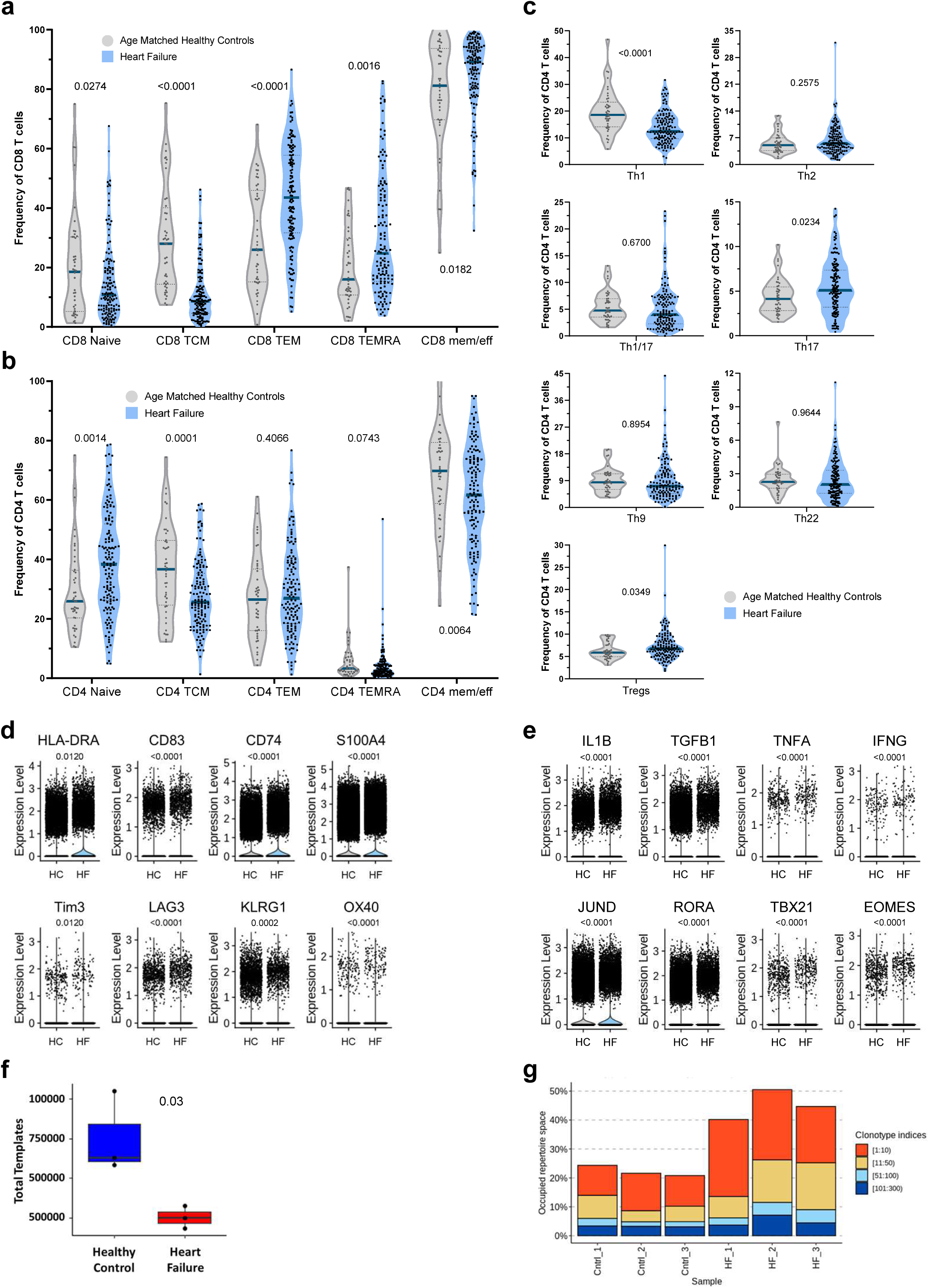
Chronic activation of T cells associates with HF. Flow cytometric analysis of (a) the CD8+ and (b) CD4+ memory and effector compartments, and (c) CD4+ helper subsets in HC (N=44) and HF (N=144) derived peripheral blood. d) Late activation and (e) cytokine gene expression profiles from scRNA-seq data of peripheral blood total T cells (N=55904 cells from N=8 HF, N=8 HC subjects). f) TCR template diversity from peripheral blood bulk TCR sequencing and (g) proportion of total TCR repertoire (i.e. occupied repertoire space) by the top 300 TCR clones in HC (N=3) and HF (N=3). Violin plots in panels a-c show median and quartiles. P-values are indicated within the graphs and were calculated via (panels a-c, f) unpaired two sided Student’s t-test or Mann-Whitney U-tests or (panels d-e) Wilcoxon Rank Sum test. TCM – T central memory; TEM – T effector memory; TEMRA – T effector memory RA+ or terminally differentiated; Th - T helper cell; Treg – T regulatory cell; HLA-DRA – human leukocyte antigen (DR-alpha chain); CD74 – Major Histocompatibility Complex, Class II Invariant Chain; CD83 – Activated B Lymphocytes, Immunoglobulin Superfamily S100A4 – S100 Calcium Binding Protein A4; Tim3 – HAVCR2; LAG3 - lymphocyte-activation gene 3; KLRG1– Killer Cell Lectin Like Receptor G1; OX40 - TNFRSF4 or Tumor necrosis factor receptor super family member 4; IL1B – Interleukin 1 beta; IFNG – Interferon gamma; TNFA – Tumor necrosis factor alpha; TGFB1 – Tissue growth factor beta unit 1; JUND – cJun delta subunit; RORA - RAR Related Orphan Receptor A; TBX21 – T-box transcription factor 21; EOMES - Eomesodermin/Tbr2

As T cell subsets strongly associated with HF status in flow cytometry data, we used scRNA-seq to gain unbiased insights into T cell signatures of T cells from circulating immune cells of HC (n=8) and HF (n=8).^12^ ScRNA-seq data showed upregulation of maturation markers (e.g. HLA-DRA, CD83, CD74) (Figure 1d). Additionally, T cell exhaustion/late activation markers like LAG3, Tim3/HAVCR2, KLRG1 were increased (Figure 1d). Corresponding regulators of exhaustion associated genes (i.e. Eomes: Eomesodermin/TBr2) that regulate Tim3 expression levels were also increased in HF T cells (Figure 1e).^13,14^ Moreover, enhanced expression of S100A4 and OX40 within HF T cells suggests that these cells show the imprint of T cell expansion and potential memory inflation (Figure 1d). Transcription factors associated with inflammation and activation were also elevated in HF T cells and include JUND, RORA and TBX21/TBET (Figure 1e). Finally, profibrotic and inflammatory cytokines (i.e. TNFA, IL1B, IFNG and TGFB1) showed significantly increased expression in HF T cells (Figure 1e).^15–18^ Overall, the scRNA-seq data confirms that T cells from HF patients are more activated with signs of exhaustion. Additionally, scRNA-seq discloses that T cells from HF patients have increased proliferation and profibrotic cytokine gene expression.

As activation of T cells results in their clonal expansion, we assessed the T cell receptor (TCR) repertoire by performing TCR sequencing (TCR-seq). Bulk TCR-seq analysis revealed a reduction of TCR template diversity within the HF derived samples (n=3) compared to age matched healthy donors (n=3) (Figure 1g). Correspondingly, the most abundant 300 TCRs made up a greater fraction of the total TCR repertoire within the HF derived T cells (Figure 1h), suggesting a greater proportion of T cells in HF respond to singular antigens.

Overall, these data imply that HF associates with chronic T cell activation, inflammation and clonal expansion.

### HF associates with enhanced T cell (self-)recruitment and homing potential

To determine whether activated and clonally expanded HF-derived T cells may have the potential to migrate to injured tissue preferentially, we assessed the expression of CCR5, which is known to mediate T cell activation and trafficking to inflamed tissue.^19,20^ We found CCR5 to be elevated within every HF derived T cell subset of the flow cytometry cohort (Figure 2a). Of note, CCR5 shows the opposite kinetics of CCR7, which is lost during maturation and whose loss enables T cells to leave the lymph node.^21,22^

**Figure 2:**
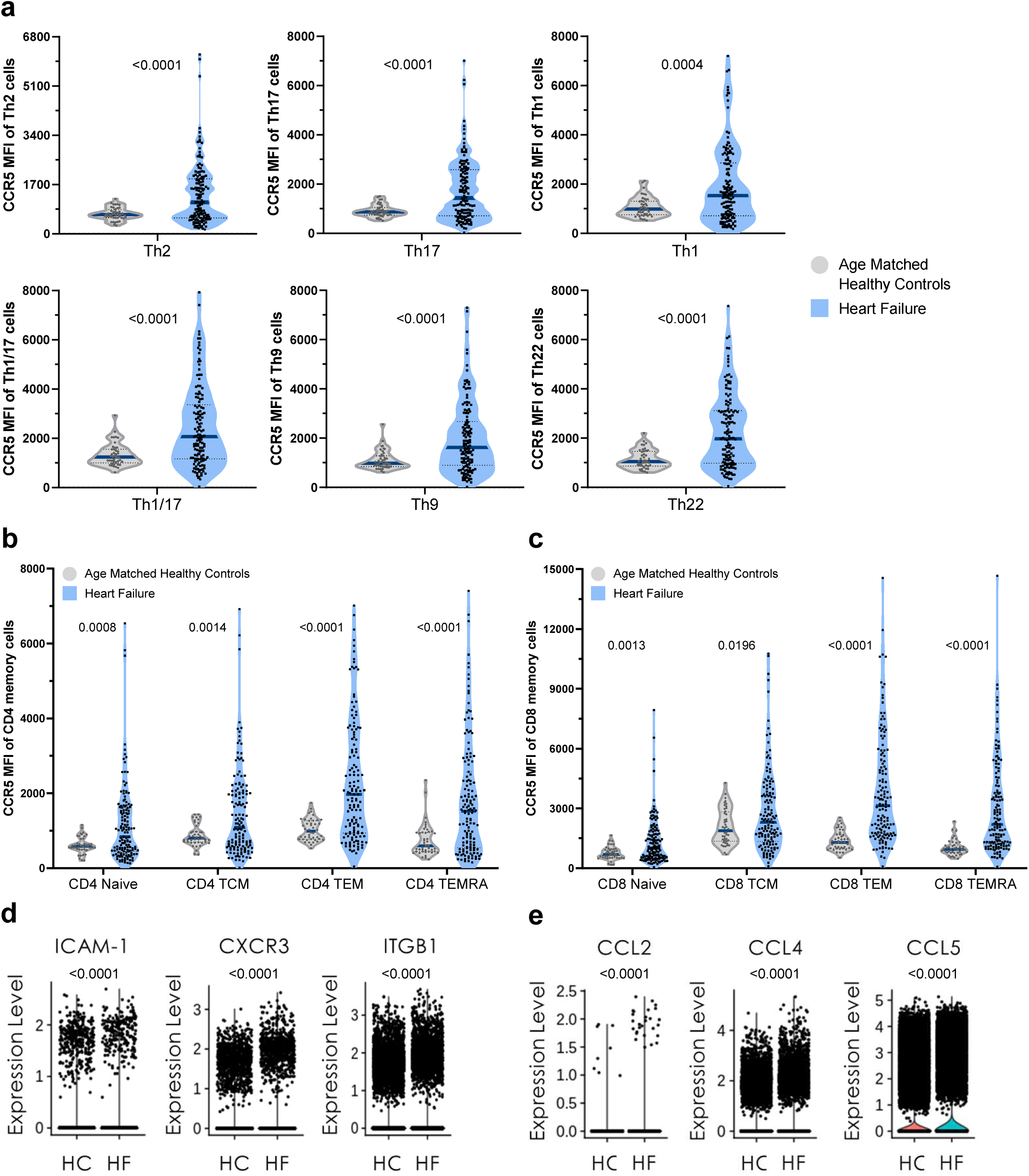
Enhanced T cell (self-)recruitment and homing potential. a-c) CCR5 expression of the peripheral blood (a) CD4+ helper subpopulations, as well as (b) CD4+ and (c) CD8+ memory populations for HF (N=139 (a) or N=144 (b+c)) and HC (N=44). Expression levels of (d) adhesion molecules/homing receptors and (e) chemokine expression levels comparing HF (N=8) and HC (N=8) from scRNA-seq analysis of peripheral blood derived total T cells. (N= 55904 cells). Violin plots in panels a-c, individual points show geometric mean of mean fluorescence intensity (MFI), with the median and quartiles being reported. P-values are indicated within the graphs and were (panels a-c) calculated via unpaired two sided Student’s t-test or (panels d-e) Wilcoxon Rank Sum test. TCM – T central memory; TEM – T effector memory; TEMRA – T effector memory RA+ or terminally differentiated T cells; Th – T helper cell; Treg – T regulatory cell; ICAM-1 – Intercellular adhesion molecule 1; ITGB1 – Integrin beta 1; CXCR3 - CXC motif chemokine receptor 3; CCL2 – C-C motif chemokine ligand 2 or monocyte chemoattractant protein-1, MCP-1; CCL4 – C-C motif chemokine ligand 4 or macrophage inflammatory protein (MIP)-1β; CCL5 – C-C motif chemokine ligand 5 or RANTES

Data from scRNA-seq confirmed this observation and showed that CXCR3 and adhesion markers such as ITGB1, ITGAX and ICAM-1 were upregulated within HF patients (Figure 2b). This observation is complemented by the elevated expression of chemoattractants like CCL5, CCL4 (ligands for CCR5) and CCL2 in T cells from HF patients. These data further support general immune cell recruitment capacity and potential “self-recruitment”/paracrine signaling of HF T cells (Figure 2c).

### Associations of clinical parameters, comorbidities and mortality with T cell populations

Association of clinical parameters and comorbidities with T cell populations was determined with a multivariate correlation and a univariate Mann-Whitney-U analysis respectively (Extended Data Tables 3 and 4). Notably we find that atrial fibrillation positively correlated with CD4^+^ TEM and TEMRA and CD8^+^ TEMRA cells, while CD4^+^ and CD8^+^ TCM frequencies (relative to total CD4^+^ and CD8^+^ T cells, respectively) inversely correlated with atrial fibrillation. Additionally, chronic kidney disease, represented by reduced GFR (glomerular filtration rate) values, was positively associated with the CCR5 surface expression levels of Th1/17 and CD8^+^ TEMRA cells. Finally, CD4^+^ TCM cells positively correlate with HDL levels and an increased GFR, indicating a possible improvement of the kidney function. Thus, specific comorbidities have distinct associations with T cell populations.

Next, we assessed if T cell subsets associate with mortality or major adverse cardiovascular events (MACE, defined by hospitalization, decompensation, second MI, cardiac surgical interventions or death) via a univariate continuous and dichotomized Cox regression analysis for death associations or a binary logistic regression analysis for associations with MACE. We observed that the mean expression of the homing marker CCR5 on CD8^+^ TEMRA and CD8^+^ Tc9 cells predicted mortality (p=0.0145 and p=0.0268, respectively), since the MFI of CCR5 on T cells of patients who later succumbed to MACE was significantly higher than in the survivor group (Figure 3a, Extended Data Table 5). Additionally, the mean frequencies of CD4^+^ TCM cells were 20.6% within the non-survival vs. 25.7% in the survival group (p=0.0636) and for CD4^+^ Tregs the frequency of the non-survival group was 4.9% and 7.0% in the survival group (p=0.0738) (Extended Data Table 5). Using the Youden index derived from the ROC curves, relating CD8^+^ TEMRA CCR5 expression and CD4^+^ TCM frequency to mortality, revealed a CD8^+^ TEMRA CCR5 expression of 4010 MFI and CD4^+^ TCM frequency of 25% as the optimal cut-off values to predict mortality during follow-up (Extended Data Figure 2b,c, Figure 4b,c). Hazard ratio (HR) for CD8^+^ TEMRA CCR5 above the cut-off value is 3.6, while patients with a frequency of CD4^+^ TCM cells below the cut-off have an HR of 16.0 (Extended Data Table 7).

**Figure 3.**
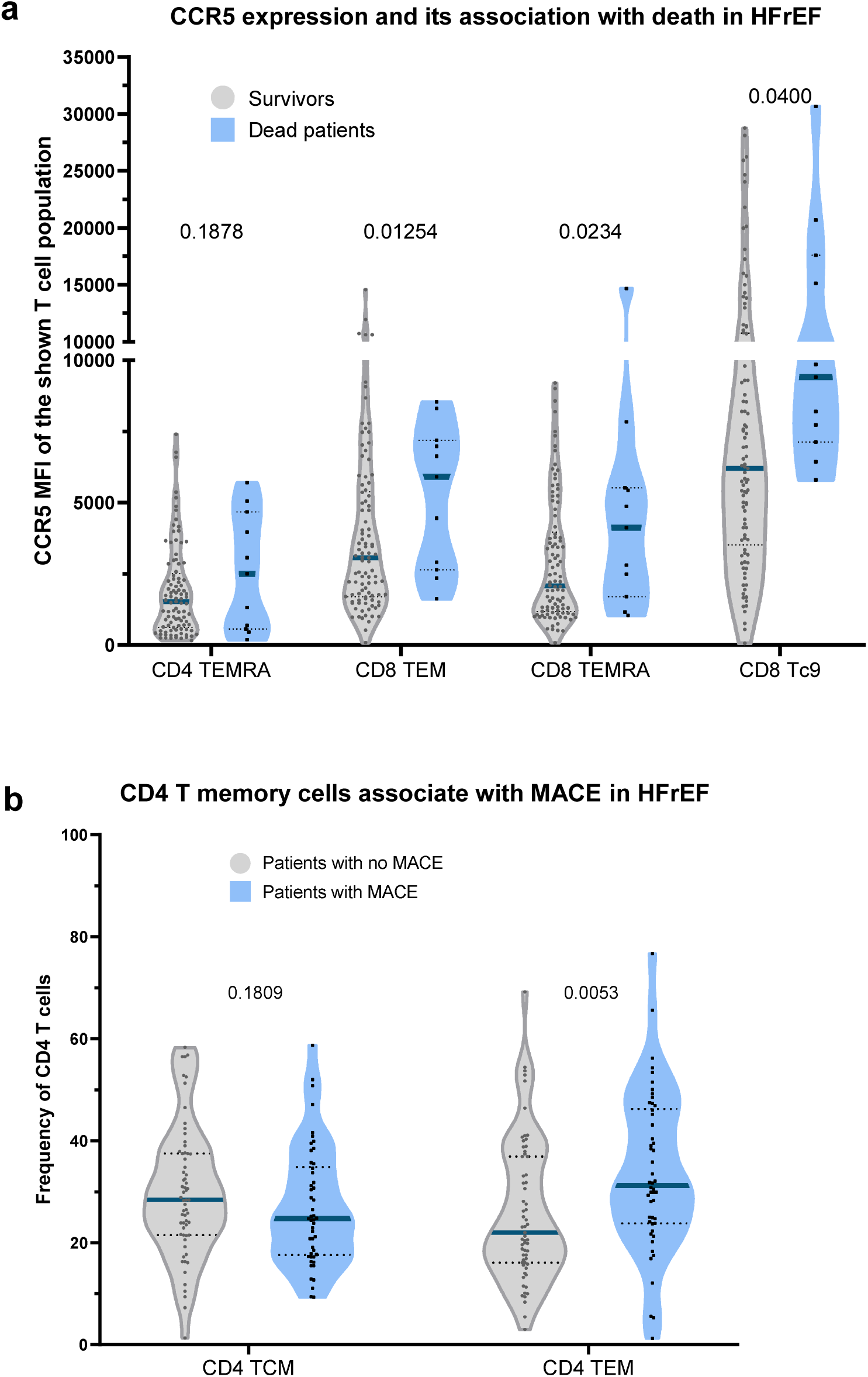
Association of immune parameters with death and MACE. a) Activation and homing/lymph node egress marker CCR5 expression was assessed across several T cell subsets within the HF cohort, comparing survivors (N=96) and dead patients (N=11). b) Frequency of CD4+ TCM and TEM cells within the HF population, comparing patients with major adverse cardiac events (i.e. second MI, re-hospitalization, cardiac surgery or death, N=50) and patients without MACE (N=63). The data are shown as violin plots with median and quartiles. P-values are indicated within the graphs and were calculated via unpaired two sided t-test. MFI – Mean fluorescence intensity; TCM – T central memory; TEM – T effector memory; TEMRA – T effector memory RA+ or terminally differentiated; Tc9 – T cytotoxic 9

**Figure 4.**
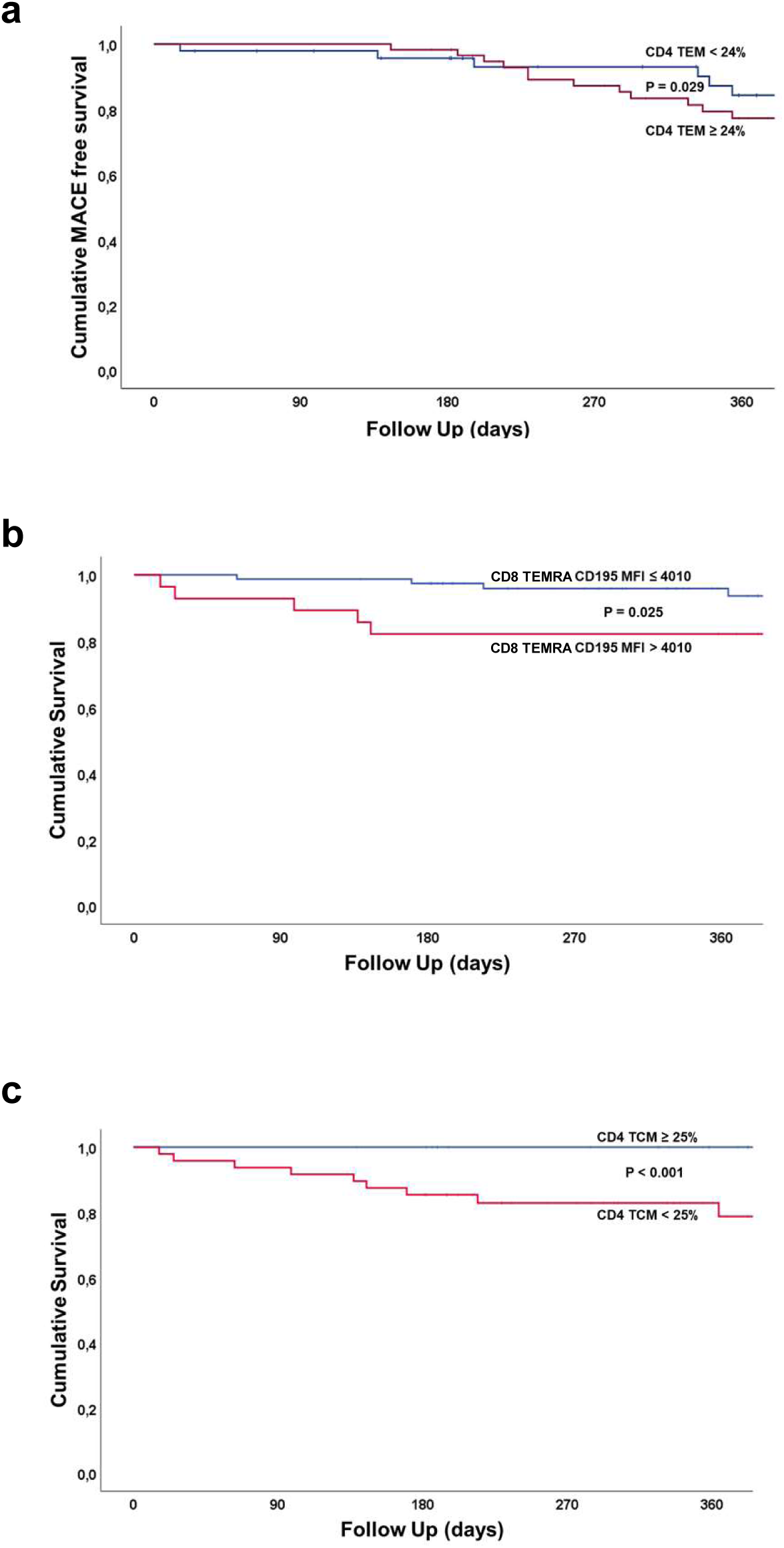
Association of immune parameters with death and MACE – correlation analysis. a) Kaplan-Meier survival curve of MACE free patients above and below 24% CD4+ TEM cells. b/c) Kaplan-Meier survival curve of cumulative survival of HF patients above and below (b) 4010 MFI of CD195 on CD8+ TEMRA cells and (c) above and below 25% CD4+ TCM cells. Population size and group size are together with the p-values indicated within the graphs and were calculated via Mann-Whitney-U test and dichotomized Cox-regression analysis. CI – Confidence Interval; MFI – Mean fluorescence intensity; TCM – T central memory; TEM – T effector memory; TEMRA – T effector memory RA+ or terminally differentiated

Although CD4^+^ TEM levels were not changed between HF and HC (Figure 1b), high CD4^+^ TEM cell frequencies associated with MACE in HF patients (HF patients without MACE: 22.3%±13.7%; HF patients with MACE: 28.9%±15.2%; p=0.0053) (Figure 3b, Extended Data Table 6). By comparison, frequencies of CD4^+^ TCM cells were 26.5% in patients without MACE vs. 24.6% in the MACE group (p=0.1809) (Figure 3b and Extended Data Table 6). Using the Youden index derived from the ROC curves relating CD4^+^ TEM cell frequency to MACE revealed a CD4^+^ TEM frequency of 24% as the optimal cut-off value to predict MACE during follow-up (Extended Data Figure 2a, Figure 4a). HF patients with a frequency of CD4^+^ TEM cells above the cut-off have an odds ratio of 3.2 (Extended Data Table 8, Supplementary Table 1). In order to assess whether the increase in MACE may be confounded by the presence of concomitant risk factors for worse outcome, multivariate analyses were performed including NT proBNP, hemoglobin, NYHA and COPD as previously established prognostic factors in HF patients. As summarized in Extended Data Table 8, CD4+ TEM > 24% remained an independent predictor with a odds ratio of 3.2 (1.4–7.2) for MACE in addition to the presence of NT proBNP.

### T cells migrate to and accumulate in the murine heart post MI

Given that mortality and MACE are predicted by increased homing and activation status of peripheral T cells, we investigated whether chronic HF leads to increased cardiac infiltration and accumulation of these T cell populations. Therefore, we employed a permanent left anterior descending (LAD) coronary artery ligation myocardial infarction in C57/BL6 mice, tracking mice over time and conducting scRNA-seq at defined intervals after MI. Analyses were conducted at days 1, 3, 7, 14, 28, and 48, with day 0 representing homeostasis (Figure 5a,b). The T cell population in the infarcted heart exhibited anticipated acute MI immune cell kinetics, with an initial increase in T cell numbers at day 3 during the acute response, which declined at days 7 and 14. Interestingly, the abundance of T cells increased again starting at day 28 and reaching levels at day 48 comparable to the frequencies of day 3 (Figure 5c). Moreover, murine cardiac T cells displayed increasing expression of Ccl4, Ccl5, and Cxcl10 over time (Figure 5d), mirroring the upregulation of T cell homing markers observed in the blood of human HF patients.

**Figure 5.**
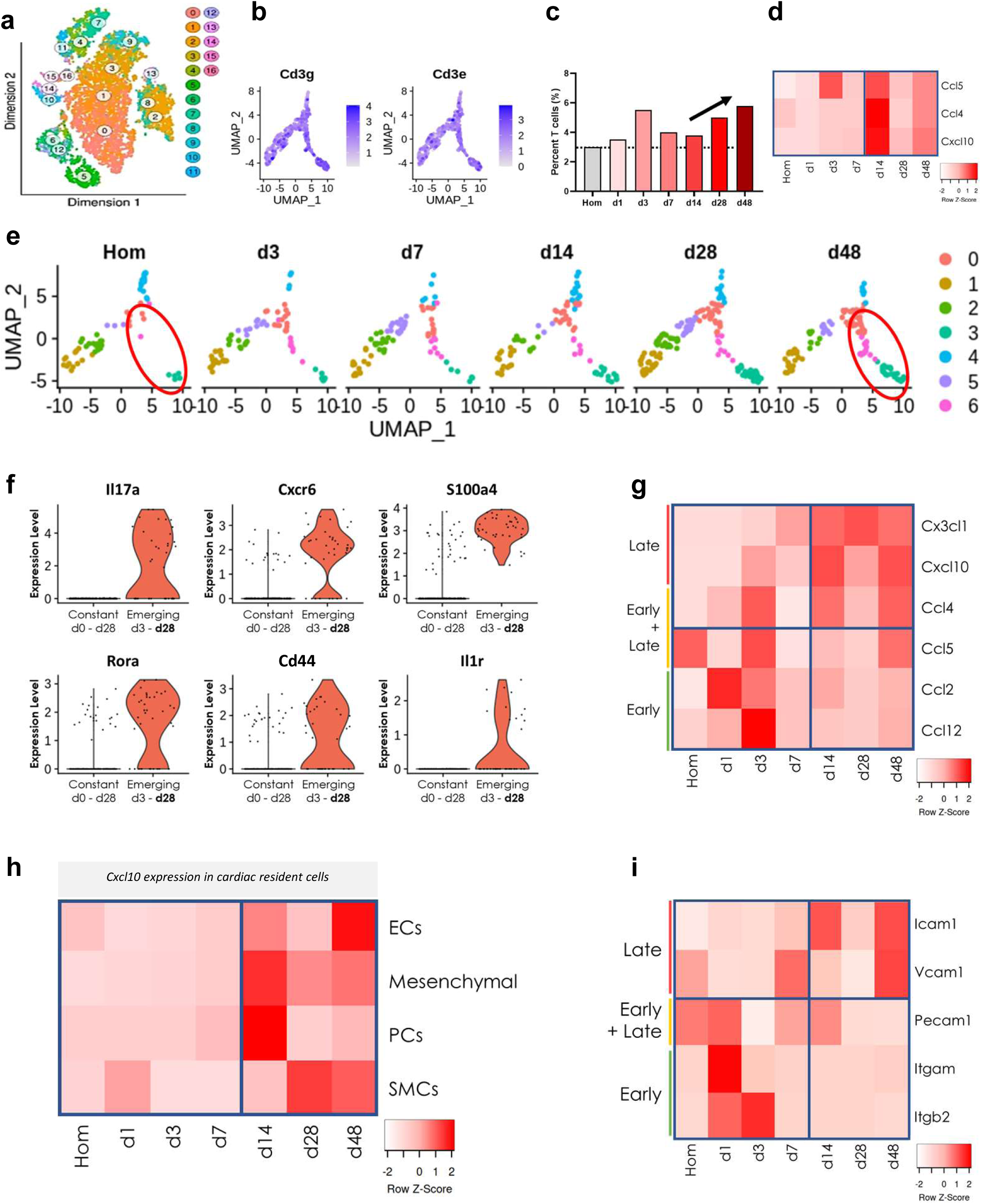
T cells migrate to and accumulate in the heart post-MI. a) UMAP plot shows clustering of all cardiomyocyte-depleted cardiac-derived cells and time points from permanent ligation model MI-time course experiment, with analyses performed at days (d) 0 (homeostasis/Hom), d1, d3, d7, d14, d28, d48. b) UMAP of T cells from all time points post MI with relative expression shown for Cd3g and Cd3e. c) Relative frequency of T cells within the heart at different time points (N=1 mouse per time point), and (d) heatmap showing mean chemokine expression levels of total T cell population at different time points. e) UMAP plot showing changes of T cell subclusters proportions over time. Late emerging clusters highlighted by red circle. f) Comparison of emerging T cell clusters with the cells from the already existing T cell clusters for the indicated genes above the violin plots. g) Relative mean chemokine expression within all cardiomyocyte-depleted cardiac-derived cells (N=9198) by time point. h) Heatmap expression patterns by cell type for cardiac-resident cells for Cxcl10. i) Mean adhesion molecule expression patterns by heatmap are shown within the endothelial population, organized by time points. N= 7 mice were harvested and 13058 cells for all mice. Statistical analysis was performed with Wilcoxon Rank Sum test and for comparing the emerging T cell clusters with the existing T cells in that UMAP location an unpaired Student’s t-test was performed. EC - endothelial cell; PC – Pericytes; SMCs – smooth muscles cells; Cx3Cl1 – Fractalkine or C-X-C motif chemokine; CxCl10 – C-X-C motif chemokine ligand 10; Ccl12 – C-C motif chemokine ligand 12; CD44 – H-CAM or Hyaluronate Receptor; Il17a – Interleukin 17 alpha; CxCr6 – C-X-C motif chemokine receptor 6; Vcam1 - Vascular cell adhesion molecule 1; Pecam1 - Platelet And Endothelial Cell Adhesion Molecule 1; Itgam1 - Integrin Subunit Alpha M

Interestingly, the post-MI T cell population underwent a phenotypic shift over the time course, as shown via dimensional reduction analysis wherein T cells formed novel clusters at late time points (Figure 5e). Comparative analysis with constant T cell clusters 0, 1, 2, 4, and 5 unveiled a highly activated (Rora, Cd44) and proliferative (S100a4) Th17 or Th1/17-like phenotype (Il17a, Cxcr6) (Figure 5f).

Investigating the non-immune, cardiac cell population (i.e. endothelial cells, fibroblasts, smooth muscle cells, pericytes, Schwann cells), we observed a temporal signature in chemoattractant profiles. The acute phase was characterized by elevated Ccl12 and Ccl2 transcript levels (Figure 5g). At day 7, Ccl5, Ccl12, and Ccl2 were downregulated. From day 14 onwards T cell homing/attraction ligands Ccl4 and Ccl5 were re-expressed. By day 48 post-MI, Cxcl10 and Cx3cl1 were increasingly expressed. Especially the subanalysis of the Cxcl10 expression pattern in the cardiac resident cells (excluding leukocytes), revealed that smooth muscle cells and endothelial cells upregulated their expression of Cxcl10 in the late stage, being the cell types that are potentially exposed the most to infiltrating and homing T cells (Figure 5h).

Subanalysis of endothelial cells showed increased expression levels of adhesion markers Icam1 and Vcam1 in the late phase post-MI (Figure 5i). This implies that endothelial cells exhibit an increased potential to allow immune cell adhesion and tissue infiltration late post-MI, substantiating a framework for chronic tissue damage through persistent low-grade immune activity and influx.

Supporting this observation of an immune cell attracting phenotypic shift within the cardiac landscape post-MI, we performed a subanalysis of spatial transcriptomic sequencing data of the human infarcted heart at different time points after MI.^23^ Within this experimental setup cardiac sections of the ischemic, border and remote zone, with the fibrotic zone representing the border zone area after 30 days post-MI, were used for analysis. Cardiac biopsies from N=19 donors at 1 to 150 days after MI were taken and compared to non-infarcted human heart sections (N=4 donors) (Extended Data Figure 3a). Importantly, data show T cell markers (e.g. CD8, IL7R) colocalize with the ischemic zone (Extended Data Figure 3c). Also, the molecular niche 3 population, defined in part by T cell markers is notably enriched in the fibrotic zone in the late stage after MI (Extended Data Figure 3b,d).

To establish potential functional impacts of the afore mentioned sustained inflammation in the post-MI the cardiac landscape and ongoing immune cell infiltration, we investigated the effects of latent inflammation on the cardiac resident cells. Thus PBMCs (comprised of ∼45-70% T cells) from HF patients and HCs were cultured for 72h and their supernatant was harvested, then applied to endothelial cells in culture. After 48h of coculture, cell viability was assessed via flow cytometry (Figure 6a). Here, HF derived supernatant induces increased apoptosis and necrosis of endothelial cells (n=4-6 per treatment, Figure 6b). Additionally, qPCR analysis showed that HF supernatant treated endothelial cells have diminished expression of the checkpoint molecule PDL1. PDL1 regulates and stops T cell responses, suggesting loss of endothelial PDL1 signaling contributes to ongoing cytotoxic activity of T cells infiltrating the heart (Figure 6c).

**Figure 6.**
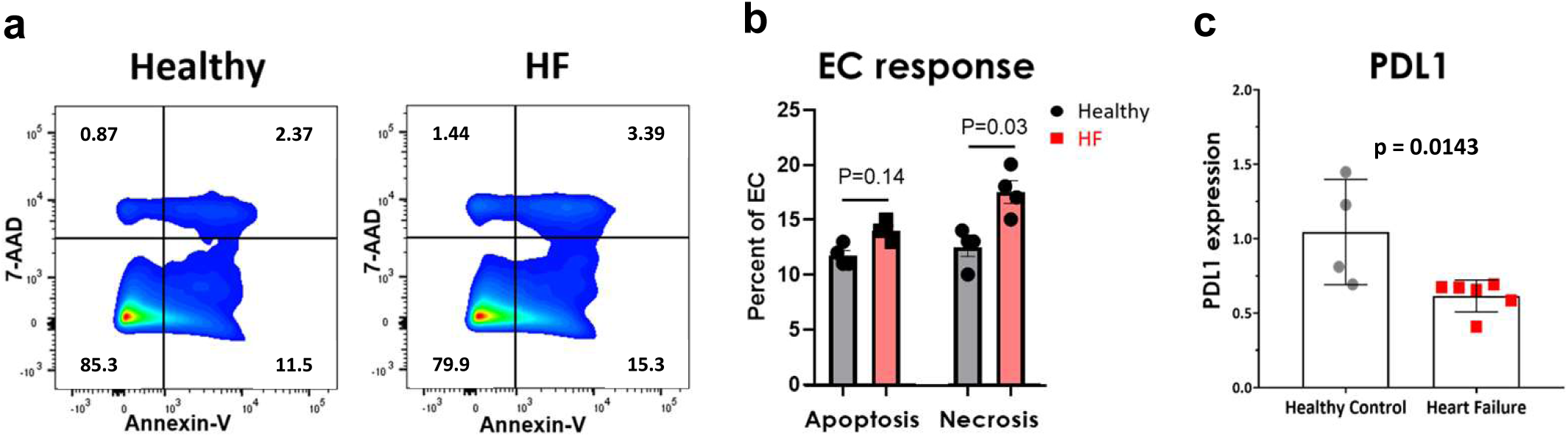
Cytotoxicity of immune cell derived SN was assessed via an Annexin-V/7-AAD staining of human umbilical vascular endothelial cells (HUVECs) after 72h. a) Exemplary flow cytometry plot for HUVECs treated with immune SN derived from HC and HF cells. b) Apoptotic and necrotic cell frequency (N=4 independent experiments). c) PDL1 transcript expression measured in HUVECs following immune SN treatment from HF cells (N=6 donors, N=3 independent experiments) and HC cells (N=4 donors, N=3 independent experiments) with the standard error of the mean (SEM) shown. P-values are indicated within the graphs and were calculated via unpaired two sided Student’s t-test. EC – endothelial cell = HUVEC; PDL1 - Programmed cell death ligand-1

### T cells clonally expand in the human heart relative to peripheral blood in HF

Having shown clonal expansion in the periphery and cardiac accumulation of T cells, we investigated whether discovered peripheral blood clones could be found in cardiac tissue. Therefore, we performed scRNA/TCR-seq from CD45^+^ cells isolated from cardiac tissue or peripheral blood from HF with comparison to peripheral blood T cells of healthy donors (Figure 7a,b). These data agree with the previous observations of the bulk TCR sequencing, showing that the peripheral blood T cells of the HF patient have relatively expanded TCR clones when compared to the four healthy donor blood samples (Figure 7c). Additionally, the HF derived cardiac biopsy TCR clonotypes could also be found within the peripheral blood T cells of the same patient, suggesting they are of peripheral origin (Figure 7d).

**Figure 7.**
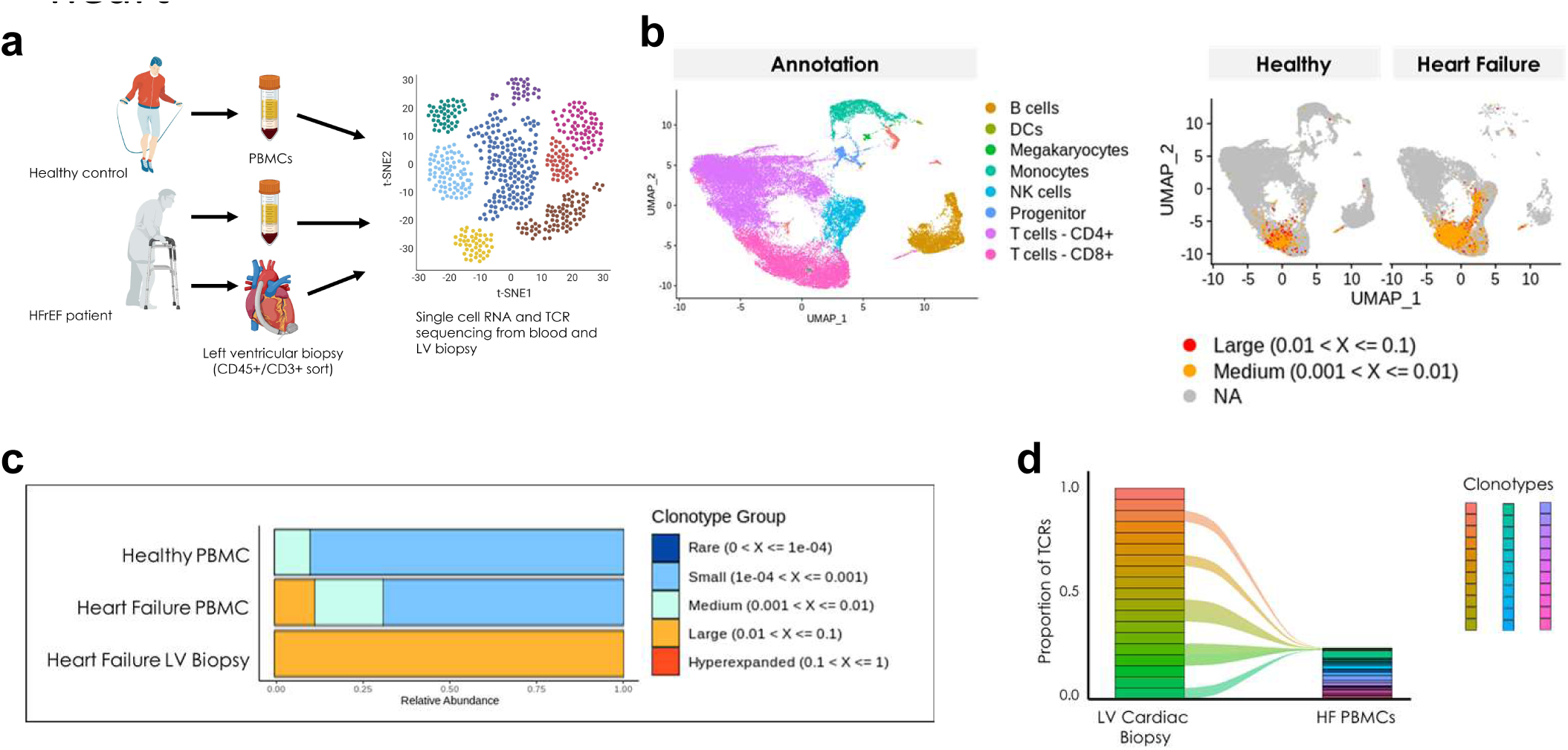
T cell clones of the PB infiltrate and expand within the heart. Schematic a) describes the general experimental/study design. PBMCs from peripheral blood and a heart-biopsy from a Left ventricular (LV) assist devised HFrEF patient were extracted to obtain total leukocytes. These cells were then used for combined single cell RNA sequencing and TCR sequencing and compared to N=4 individual healthy derived whole blood isolated PBMCs. b) gives the annotation of the PBMCs found within all combined samples after using dimensional reduction and UMAP. An overlay of the identified TCRs with their barcodes fits with the CD4 and CD8 T cell clusters. The general relative abundance of the TCR clones found within the combined healthy controls and the HF derived PBMCs and Biopsy are shown in graph c). The graph d) shows that all of the HF biopsy derived TCRs can be also found within the TCR repertoire of the same patients peripheral blood. Analysis of N=26241 total T cells was performed with scRepertoire.

To finally evaluate which antigens are targeted by TCRs, we utilized epitope prediction modeling tools (i.e. TCRMatch) analyzing the previously mentioned bulk TCR sequencing dataset (Extended Data Figure 4a). For this purpose, we compared epitope prediction across all donors (Extended Data Figure 4b). With this information we could identify epitopes belonging to 23 proteins of human origin that were only associated with the TCRs from HF patients. Of note, some proteins have high sequence similarity to those restricted to the myocardium (i.e. Ryanodine Receptor 3). Thus, we revealed a potential set of HF-restricted TCRs with autoimmune capacity (Extended Data Figure 4c).

## Discussion

Our multi-modal study employing gold standard flow cytometry and scRNA-seq techniques presents groundbreaking insights into T cell modulation in HF. Specifically, we demonstrate that proinflammatory CD4^+^ Th17 cells, CD8 TEM and TEMRA cells along with expression of the homing marker CCR5 on T cell subsets positively associate with HF. Strikingly, CD4^+^ TEM cells associate with MACE while CCR5 expression on CD8^+^ TEMRA cells positively associate with mortality in HF patients. TCR sequencing demonstrates that HF patients have increased clonal expansion of T cells in the peripheral blood and heart relative to healthy controls. Importantly, an unbiased TCR repertoire analysis provides a list of novel HF patient-restricted protein targets. These data suggest that T cells not only predict mortality but that they have increased probability of autoreactivity.

This study highlights T cell dysregulation in chronic heart failure. Building on prior studies, we observe dysfunctional and proinflammatory T cells that have been implicated in driving adverse cardiac remodeling.^2,24–26^ Notably, a shift in Tregs towards a Th17-like phenotype is identified in peripheral blood, suggesting Tregs may lose their immunosuppressive capacity. Additionally, the post-infarcted cardiac tissue harbors an entirely novel pro-inflammatory T cell subset, potentially contributing to pathology.^27,28^ These cells show higher expression levels of activation and proliferation markers such as Cd44 and S100a4, as well as a pro-inflammatory Th1/17-like phenotype defined by high expression of Jund, Il17a and Rora.

Exploring further the impact of different immune compartments and their relation to a patient’s comorbidities we find that relative higher frequencies in circulating memory T cells associate with fewer comorbidities and decreased mortality. However, increases in CD4^+^ TEM cells positively correlated with major adverse cardiovascular events (MACE). Strikingly, CCR5 expression on CD8^+^ TEMRA cells positively correlates with mortality in HF patients. TEM cells are direct mediators of immune responses with greater homing potential (e.g. tissue specificity, tissue entry). More immune mediators in the periphery could result in greater T cell infiltration and damage to tissues, suggested by the association with MACE in HF patients. When relative abundance of TCM populations (CD4^+^ and CD8^+^) is compared between age-matched HC and HF patients, TCM frequency is nearly halved in HF patients. This study suggests that comorbidities, HF and mortality associate with a loss of TCM cells. To what extent decreased TCM cells may promote loss of homeostasis and protection requires further investigation.

Elucidating the molecular landscape and inflammatory environment in chronic HF, we used ScRNA-seq analyses in humans and mice. We highlight a persistent activation phenotype, increased exhaustion markers, and a Th1 and Th17-like phenotype in humans and mice with chronic HF. Elevated cytokine expression underscores the pro-inflammatory environment in HF, potentially contributing to tissue damage and fibrosis. The persistent activation phenotype of T cells in HF patients was further characterized by increased expression of mature T cell markers (CD83, CD74) and increased expression of exhaustion/late activation markers (Tim3, HLA-DRA, etc.). Transcription factor analysis supported these observations, indicating prolonged inflammation and a Th17-like phenotype in T cells. Importantly, Th17 cells were correspondingly increased in the circulation of patients with ischemic heart disease and diffuse cardiac remodeling (or fibrosis).^29^ Cytokine expression profiles and serum analysis further emphasizes the pro-inflammatory environment in HF, with elevated levels of inflammatory genes (RORA, JUND) and cytokines (TNFA, IFNG and TGFB), potentially contributing to tissue damage and fibrosis.

This study also addressed the migratory behavior of T cells in chronic HF. Elevated expression of homing markers, such as CCR5, CXCR3, and adhesion molecules, indicated enhanced homing potential of T cells in HF patients. Strikingly, CCR5 expression on CD8^+^ TEMRA cells was significantly elevated in patients with a poor prognosis in comparison to survivors of the studied HF cohort. Even the global association of CCR5 on nearly every T cell subset showed significant correlations with hyperuricemia and chronic kidney disease. These data are supported by previous murine studies, wherein a CCL5 neutralizing antibody reduced MI scar size and reduced chemokine levels in the circulation.^30^ However, this murine study did not analyze which immune cell might be implicated in this process. Additionally, the upregulation of chemo-attractants in peripheral T cells as well as murine cardiac cells suggested a coordinated immune cell recruitment mechanism, potentially contributing to sustained inflammation and tissue damage.

Our investigation extends beyond peripheral blood, employing scRNA-seq analysis of cardiac tissue to unveil clonally expanded T cells potentially contributing to chronic tissue damage. Spatial transcriptomic sequencing in the infarcted human heart highlights the localization of T cells in the fibrotic zone 30 to 150 days after MI, emphasizing the need for further exploration in maladaptive cardiac remodeling.^23^

Unbiased T cell receptor (TCR) repertoire analysis reveals novel putative targets found exclusively in clonally expanded T cell populations of HF patients. TCRMatch epitope prediction modeling revealed unique epitopes/proteins associated with T cells from HF patients, such as Ryanodine Receptor 3, which has high similarity to cardiac-restricted Ryanodine Receptor 2. This suggests a specific immune signature that may contribute to prolonged inflammation and tissue damage.^31^ Our data suggests causality of T cell interactions in cardiovascular disease development and prognosis, aligning with existing literature.

Considering translational perspectives, the findings support the potential benefits of blocking T cell activation in HF, as demonstrated by the FDA-approved drug abatacept.^7,32,33^ Ancillary findings from clinical trials (i.e. abatacept in rheumatoid arthritis vs. TNF inhibitor treatment) support the model that blocking T cell activation is particularly beneficial in mitigating MI and the need for coronary revascularization.^34^

Targeted therapies for the adaptive immune system, including T cell-mediated vaccine therapies, offer promising avenues for future research and clinical trials.^35^ Recent reports have indicated that T cell responses post-MI may have cardiac myosin targeting potential.^36,37^ While, these studies were important, they were found to be HLA restricted (minor portion of HF population) and derived from a pre-defined screen. Thus, using unbiased screens of epitopes targeted by HF-restricted inflated T cell clones may allow for more precise treatment of ischemic HF.

In conclusion, our study provides a comprehensive understanding of T cell involvement in chronic HF, shedding light on the intricate interplay between immune responses, T cell activation, and homing. These data reveal the immunological fingerprint of HF is a prematurely aged immune system. With this knowledge, our study offers new therapeutic targets that could mitigate chronic inflammation and improve patient outcomes in a substantial proportion of the HF population. Further research is crucial to explore the translational implications of our observations and to develop targeted interventions for modulating the immune response in chronic HF.

## Material and methods

### Aged Healthy Controls included within this study

Aged healthy controls included within this study were gathered via two sources. First was via buffy coats from whole blood donations (n=29 donors) to German Red Cross Blood Service Baden-Württemberg-Hessen. Donor health was ascertained by donor physicians using internationally agreed blood donor eligibility criteria supported by questionnaires developed by the national body for hemotherapy. Buffy coats were diluted 1:5 for flow cytometry analyses. The second source of healthy donors (n=15 donors) came from the staff members of the Department of Nephrology, Goethe University hospital (Frankfurt, Germany). Volunteers were excluded if having known auto-immune, heart or kidney diseases. Subsequently, 15 mL EDTA blood was drawn and further processed without dilution. Ischaemic heart failure patients with a reduced ejection fraction (LVEF<45%) were recruited by the Department of Cardiology, University Clinics (Frankfurt, Germany). Blood was obtained from these patients at approximately the same time (between 10:00 and 12:00) on study days. Informed consent was obtained from all patients. The study was approved by the local ethics review board and complies with the Declaration of Helsinki. Patients were eligible for inclusion into the study if they had stable chronic heart failure symptoms, New York Heart Association (NYHA) classification of at least II and had a previous myocardial infarction at least 3 months before recruitment. Exclusion criteria were the presence of acutely decompensated heart failure with NYHA class IV, an acute ischemic event within 3 months before inclusion, a history of severe chronic disease, documented cancer within the preceding 5 years or unwillingness to participate. Blood for the flow cytometer measurement was collected in 15 mL EDTA blood tubes, while plasma was obtained from whole blood in sodium citrate–containing cell preparation tubes (CPT Vacutainer, Becton Dickinson), which were centrifuged at 1,500g for 15 min at room temperature. Also, here EDTA blood was processed without dilution.

Ethical approval was received from the local ethics committees at each of the research centers. All participants provided written informed consent and were medically screened before participation. Samples were gathered under the ethic vote KardioBMB#2021003 Folgeantrag#1 and #2 and the votum #329/10 - “anonymized use of Buffy Coat from whole blood donations” respectively.

### PBMC isolation

Mononuclear cells from peripheral blood comprising lymphocytes and monocytes were isolated by density gradient centrifugation using Ficoll Paque (d=1.077 g/ml). Peripheral blood was distributed into 50 mL conical tubes and diluted to a volume of 35 mL with PBS (Thermo Fisher 14190144). The diluted blood was layered on 15 mL Ficoll/Percoll solution (PAN-Biotech P04-63500) without mixing the two phases.

The Ficoll layered samples were transferred to a swing bucket centrifuge and centrifuged at 800 g for 20 min at room temperature. The brakes’ de-accelerating speed was set to 0.

The white interphase between the suspension and Ficoll solution, carrying the mononuclear leukocytes, was carefully transferred to a fresh 50 mL conical tube and filled up to 50 mL with PBS. The sample was then centrifuged at 800 g for 10 min at RT with the full brake speed.

Afterwards the supernatant is removed and the pellet again resuspended in 50 mL PBS, the centrifugation step (800 g 10 min RT) was repeated. After resuspending the PBMCs a second time, an aliquot for counting was taken while the sample was centrifuged for the last time before the PBMCs (≥5×10^7^ cells/ml) were frozen for future experiments in DMSO and FCS containing medium (40% FCS heat inactivated, 50% RPMI1640 and 10% DMSO).

### Flow cytometric analyses

Multi-color flow cytometry was performed to analyze a panel of T cell surface markers. For this purpose, 60 µL of the sample (undiluted EDTA blood or 1:5 diluted buffy coat) were transferred into a 5 mL round bottom Falcon tube (FACS tube) and 37.5 µL antibody-cocktail (resulting in a 1:20 or 1:50 working dilution for each individual antibody; for antibodies included see Extended Data Table 9) was added to the sample. The full-stain sample tube is vortexed and incubated at room temperature (RT) for 20 min in darkness. Additionally, one unstained sample tube for comparison, without any antibodies or viability dye, is prepared and treated the same way as the full-stained tube. After incubation 2 mL 1x RBC-Lysis Buffer (Biolegend 420302) were added and the samples carefully vortexed. Again, the samples were incubated for 15 min at RT in darkness, afterwards 2 mL Cell Staining Buffer (Biolegend 420201) were added on top for washing. Following centrifugation at 500 g for 5 min at 4°C, the supernatant was afterwards removed and the pellet in 2 mL Cell Staining Buffer resuspended. This washing step was repeated and the final pellet was resuspended in 400 µL Cell Staining Buffer. Next 4 µL 7-AAD (Biolegend 420404) were added to the full-stain sample and vortexed. The samples were then incubated for another 10 min at RT in darkness and analyzed via flow cytometry (BD Fortessa LSR X-20). Assessed samples were analyzed in FlowJo v.10.1 applying the gating strategy shown in Extended Data Figure 5.

### Cytokine and secretome analysis

A customized 13-plex Legend plex assay was generated via Biolegend to detect the following markers: CX3CL1, CCL2, sICAM-1, TNFSR-I and TNFSR-II, IL6, IL1β, IFNy, TGFβ, TNFα and Granzyme B. HF and HC derived serum samples were diluted 1:2 and 1:100 for ICAM1 measurements via the LEGEND-plex assay. The assay was performed following the manufacturer’s (Biolegend) standard procedure in order to assess the serum concentrations. Samples were measured in duplicates and the assay was split into a 12-plex run, without ICAM-1 detection beads and into a 1-plex run for sICAM-1 due to the different pre-dilution factors needed. Assessment of samples was performed according to the standard protocol on a BD Fortessa LSR X-20 flow cytometer using the HTS (high throughput sample) plate reader to automatically acquire samples from a 96-well plate.

### Cell culture media

#### PBMC culture medium

General culture medium for isolated PBMCs is prepared using RPMI 1640 Glutamax (Thermo Fisher 61870010) with 10 mM HEPES (Thermo Fisher 15630056), 20 µM Beta-Mercaptoethanol (Thermo Fisher 31350-10), 1x sodium-pyruvate (Thermo Fisher 11360039), 10% FCS (heat inactivated), 5 ng/mL M-CSF (Peprotech) and 1x Penicillin/Streptavidin (Thermo Fisher).

#### Endothelial growth medium

EBM-2 Basal Medium (Lonza CC-3156) was supplemented with EGM-2 SingleQuots (Lonza CC-4176) as well as 1x Penicillin and Streptavidin (Thermo Fisher) and 10% FCS.

#### THP1 culture medium

THP1 monocytes were cultured in RPMI 1640 Glutamax (Thermo Fisher 61870010) with 10 mM HEPES (Thermo Fisher 15630056), 20 µM Beta-Mercaptoethanol (Thermo Fisher 31350-10), 1x sodium-pyruvate (Thermo Fisher 11360039), 10% FCS (heat inactivated) and 1x Penicillin/Streptavidin (Thermo Fisher).

### Functional assays for immune cell secretome on target cells

To study the effects of the immune secretome, PBMCs from healthy donors (HC) and HF patients were thawed and afterwards placed in 5 mL PBMC culture medium at 37°C for 72 h. The cells were daily vortexed carefully within the solution. After the incubation, the cells were pelleted (spin at 500 g for 5 min) and the supernatant taken as immune cell secretome. This secretome was then applied to HUVECs in culture in a 1:5 ratio with their normal culture medium (endothelial growth medium). Comparing HC and HF derived immune secretome effects on these cells by simultaneously treating cells with either of these supernatants. After 48 h of culture the cells were used for RNA isolation in order to track inflammation associated markers, or were trypsinized (TrypLE Select Enzyme, Thermo Fisher, 12563029) and stained for Annexin-V (BV421, Biolegend, 640924 and Annexin V Binding Buffer 422201) and 7-AAD, following the manufacturers protocol (Biolegend), in order to assess cell viability.

### ScRNA-seq study population and blood collection

Blood and cardiac biopsy were obtained from chronic heart failure patients receiving a left ventricular assist device following approval by the Ethics Committee of the Charité-University Medicine Berlin (EA4/028/12 and EA2/283/20) with written informed consent of the patients as complies with the Declaration of Helsinki. Patients were eligible for inclusion into the study if they had chronic heart failure symptoms New York Heart Association (NYHA) classification of at least III and had a previous myocardial infarction at least 3 months before recruitment. Exclusion criteria were the presence of a history of severe chronic disease, documented cancer within the preceding 5 years or unwillingness to participate. Blood was collected in sodium citrate–containing cell preparation tubes (CPT Vacutainer, Becton Dickinson), which were centrifuged at 1,500g for 20 min at room temperature. The upper layer containing mononuclear cells and plasma was collected, and cells were washed and then stored until used for droplet scRNA-seq. The left ventricular cardiac biopsies were washed, placed in a 10% DMSO freezing media and frozen at −80°C in a CoolCell container with PBMCs and stored until used.

Peripheral blood scRNA-seq data (Figures 1d,e and 2d,e) are derived from previously published data as indicated.

Peripheral blood scRNA-seq immune profiling data (n=4) (Figure 6a-d) are derived from previously published data as indicated.

### Peripheral blood and cardiac biopsy sample preparation

Frozen PBMCs were thawed in a pre-heated water bath at 37°C for two minutes and transferred into 25 ml RPMI medium followed by centrifugation at 400 g for 5 min. Then PBMCs were washed twice with PBS containing 0.04% BSA performing centrifugation steps at 200 g for 10 min. An aliquot was stained with 0.04% trypan blue to microscopically inspected and count the cells using a Neubauer chamber. Cell suspensions containing ≥ 20% dead cells after thawing were further processed by depletion of dead cells using the Dead Cell Removal Kit (Miltenyi 130-090-101). Finally, viable cells were adjusted to a cell concentration of 1000 cells/µl before undergoing scRNA-seq library preparation.

Cryopreserved cardiac biopsies were thawed on ice and minced into small cubical pieces using surgical scissors. Additionally, the tissue was dissociated by enzymatic digestion with a mixture of 500 U/ml Collagenase Type I (C0130-100MG), 1000 U/ml Collagenase Type XI (C7657-25MG), 60 U/ml Hyaluronidase (H1115000), 60 U/ml DNAse I (D5307-1KU) and 20 mM HEPES (all from Sigma Aldrich) in 1 ml PBS for 45 min at 37°C. Cell suspensions were washed with PBS containing 0.5% BSA followed by filtration through a 70 µm and 40 µm Flowmi cell strainer. Afterwards, cell suspensions were depleted from dead cells using the Dead Cell Removal Kit (Miltenyi 130-090-101) according to the manufacturer’s instructions. The remaining viable cells were stained with CD45-BV510 (Biolegend 368526) to enrich CD45-positive cells by cell sorting on a FACS Aria Fusion (BD). Sorted CD45-positive cells were washed with PBS containing 0,04% BSA and immediately subjected to scRNA-seq and scTCR-seq library preparation.

### scRNA-seq and scTCR-seq library preparations

Cellular suspensions were loaded on a 10x Chromium Controller (10x Genomics) according to manufacturer’s protocol based on the 10x Genomics proprietary technology. scRNA-seq libraries were prepared using Chromium Single Cell 5′ Reagent Kit (10x Genomics) according to the manufacturer’s protocol. In brief, the initial step consisted of performing an emulsion capture where individual cells were isolated into droplets together with gel beads coated with unique primers bearing 10x cell barcodes, unique molecular identifiers (UMIs) and poly(dT) sequences. Reverse transcription reactions were engaged to generate barcoded full-length cDNA followed by the disruption of emulsions using the recovery agent and cDNA cleanup with Dynabeads MyOne Silane Beads (Thermo Fisher Scientific). Bulk cDNA was amplified. Amplified cDNA product was cleaned with the SPRIselect Reagent Kit (Beckman Coulter). Indexed sequencing libraries were constructed using the reagents from the Chromium Single Cell 5′ Reagent Kit version 2, as follows: VDJ amplification, fragmentation, adaptor ligation; size selection with SPRIselect; with gene expression library having fragmentation, end repair and A-tailing, adaptor ligation; post-ligation cleanup with SPRIselect; sample indexing and cleanup with SPRIselect beads. Library quantification and quality assessment was performed using Bioanalyzer Agilent 2100 using a High Sensitivity DNA Chip (Agilent). Indexed libraries were equimolarly pooled and sequenced on two Illumina NovaSeq 6000 instruments using paired-end reads with sequencing read parameters being Read 1: 26 cycles; i7 index: 10 cycles; i5 index: 10 cycles; Read 2: 90 cycles.

FASTQ files were processed using Cell Ranger 7.0.0 (10x Genomics) and aligned to the human reference genome GRCh38 according to the manufacturer’s instructions.

### Gene expression analysis from scRNA-seq datasets

Analysis was performed in Seurat (version 4) R (version 3.6). Data were filtered based on the number of genes detected per cell (cells with fewer than 200 genes per cell were filtered). A global-scaling normalization method for the gene–cell–barcode matrix of the samples was employed. We normalized the feature expression measurements for each cell by the total expression, multiplied this by a scale factor (10,000) and log-transformed the result to yield the normalized unique molecular identifier (nUMI) value later reported. Regression in gene expression was performed based on the number of nUMIs. Principal component analysis (PCA) was run on the normalized gene–barcode matrix. Barnes–Hut approximation to UMAP was then performed on principal components to visualize cells in a two-dimensional space. This graph-based clustering method relies on a clustering algorithm based on shared nearest neighbor (SNN) modularity optimization. Differential transcriptional profiles by cluster were generated in Seurat with associated GO terms derived from the functional annotation tools DAVID Bioinformatics Resources 6.7 (NIAID/NIH, https://david.ncifcrf.gov/summary.jsp) and Metascape (http://metascape.org).

### Analysis of the scTCR-seq repertoire

The tool scRepertoire was primarily used for VDJ analysis of scRNA-seq data. scRepertoire was used to call clonotypes using the CDR3 amino acid/nucleotide sequences, by gene usage, and by the combination of CDR3 nucleotide sequences and genes. In brief unique clonotypes were identified, total and relative abundance of clonotypes were determined, the distribution of CDR3 nucleotide and amino acid sequences for clonotypes were assessed. Importantly, more in depth analysis of clonal architecture was performed for clonal space occupied by clonotypes of specific proportions and diversity determined across samples.

### Bulk TCR sequencing preparation and sequencing

Genomic DNA was extracted from the peripheral blood utilizing the AllPrep DNA/RNA Kit (Qiagen) and was sent to Adaptive Biotechnologies (Seattle, USA) for bulk TCRβ sequencing. Sequencing data were aligned by Adaptive Biotechnologies. Data was processed using the ImmunoSEQ Analyzer (Adaptive Biotechnologies, v3.0).

### Bulk TCR repertoire data analysis

TCR repertoire analysis was performed by integrating TCR sequences from all samples into the Immunarch 0.9.0 immunoinformatic analytic package in R (R Foundation for Statistical Computing, Vienna, Austria). Focus was placed on the TCR CDR3β regions amino acid and nucleotide sequence as it has been previously shown to play the largest part in receptor specificity. Diversity was calculated using the Hill Foundation number and Chao1 richness index.

Putative epitopes were predicted from antigens using TCRMatch, uses k-mers to compare the overlap in motifs based on the top 300 CDR3β sequences per donor to identify against similar sequences annotated in the Immune Epitope Database (IEDB). The TCR sequencing data were submitted to the TCRMatch online tool (http://tools.iedb.org/tcrmatch/) and epitopes filtered based on target species. Predicted epitopes from homo sapiens (species) were then compared.

### RNA extraction

Cell culture supernatant from wells was removed and the cells were carefully washed with PBS. Afterwards the cells were harvested by adding 700 µL Qiazol on top, followed by an incubation step with closed lid for 5 min. Then the suspension was transferred by pipetting up and down to a fresh 1.5 mL Eppendorf tube. RNA isolation followed the standard procedure using the miRNeasy Micro Kit (Qiagen 217084) according to the manufacturer’s protocol.

### cDNA synthesis

cDNA synthesis of 1000 ng RNA was performed following the standardized procedure of the manufacturers M-MLV Reverse Transcriptase protocol (Thermo Fisher 28025-013).

For 1x reaction (Rx) 1000 ng RNA was prepared diluted with H_2_O in up to 11 µL total volume. Then 1 µL random hexamers (Thermo Fisher S0142) and 1 µL dNTPs (Thermo Fisher R0191) are added and the mix is heated for 5 min at 65°C. After cooling the sample on ice, the tube is centrifuged briefly to bring down condensed water. Next, 4 µL of 5x First Strand Buffer (Thermo Fisher Y02321) are added together with 2 µL DTT 0.1 M (Thermo Fisher Y00147), and then Reverse Transcriptase (1 µL). cDNA synthesis in a Thermocycler is started afterwards.

### qPCR

qPCR assays were run in 384 well format with a reaction volume of 10 µL, technical triplicates of each sample per primer set investigated was prepared. The reaction mix consisted of 5 µL (2x) FAST SYBR Green, (Thermo Fisher 4385612) 0.25 µL of each the forward and reverse primer (10 µM), 2.5 µL H_2_O (ultrapure) and 2.5 µL cDNA template (between 4 to 5 ng/µL). A negative control containing H_2_O instead of cDNA template was included.

The following primers were used: RPLP0 (60S acidic ribosomal protein P0) with the referenced forward: ATCCGTCTCCACAGACAAGG and reverse primer: TCGACAATGGCATCTAC sequence, as well as PDL1 (Programmed cell death ligand 1) with the described forward: TGGCATTTGCTGAACGCATTT and reverse primer: AGTGCAGCCAGGTCTAATTGT sequence.

RPLP0 was used as housekeeping gene. Assays were run on a ViiA 7 instrument with Quant Studio Real-Time PCR Software (both Applied Biosystems). The cycling protocol was as the following: 95°C for 20 sec followed by 40 cycles of 95°C for 1 sec and 60°C for 20 sec, final melt curve stage (95°C 15 sec, 60°C 1 min and 95°C 15 sec). Data was analyzed using the ΔΔCt method.

### Statistics

Statistics were done using R (Seurat v4), PRISM version 8.0.1 and SPSS version 27. The sample size of each experimental group is described in the corresponding figure caption, and all of the experiments were conducted with at least three biological replicates unless otherwise indicated. GraphPad Prism was used for all statistical analyses except for sequencing-data analysis or the univariate analysis. Quantitative data displayed as histograms are expressed as mean ± s.e.m. (represented as error bars). Results from each group were averaged and used to calculate descriptive statistics. Mann–Whitney U-tests (unpaired samples, two-tailed) and Student’s t-tests (unpaired, independent samples, two-tailed) were used for comparisons between groups unless otherwise indicated. P < 0.05 was considered to be statistically significant. Experiments were not randomized.

Associations of clinical parameter and comorbidities with certain immune component changes was executed using Mann-Whitney-U-Test, ROC-Curve-Analysis and Kaplan-Meier-Analysis with log-rank-test and Chi squared-test, where appropriate. The differences were considered significant by error probability P<0.05. In detail: For multivariate analysis the clinical parameters were compared using multivariate correlation analysis, while the sub-analysis for death and MACE associations with the immune components was done in the following way: Subsets or markers were tested via Student’s t-tests (unpaired, independent samples, two-tailed) by splitting patients into alive and dead or into patients with MACE vs patients without MACE. Additionally, after ROC-Curve Analysis and Youden-Index, a univariate continuous and dichotomized Cox regression analysis was performed for the mortality associations. A continuous and dichotomized binary logistic regression analysis was performed for MACE. Significant associations within the mortality and MACE subgroups were used to find a Youden-Index based cut-off value for a dichotomized based Kaplan-Meier-Analysis with log-rank-test. For the MACE subgroup, the significant parameters of the univariate analysis were used for a multivariate analysis.

## Acknowledgments

We want to express special gratitude to the Institute for Transfusion Medicine and Immunohematology (Frankfurt, Germany), Departments of Cardiology and Nephrology, Goethe University Hospital (Frankfurt, Germany) for their contributions and Anje Müller, Katrin Krüger and Heike Meyborg, Deutsches Herzzentrum der Charité (Berlin, Germany) for their technical assistance. This work was supported by Cardiopulmonary Institute Excellence Cluster, the ENABLE Consortium and DFG funded SFB1531 Research Consortium. SC was supported by the DFG (SFB 1531, Project number 456687919; Project B10).

## Author Contributions

MM, WTA and SD developed the theory, outlined the project idea and planned the experiments. The experiments and data shown were carried out by MM, TR, LT, BS, KH, EM IM, and WTA. SC, CK, TS, DL, HB, AMZ and TS supervised ethics approvals and for the human study and/or acquire blood from HF patients and healthy volunteers. AB, DJ, LT, MM and WTA analyzed the data. MM, WTA, EU, AMZ and SD supervised and verified the analytical methods. AMZ, EU, SD, HB, TS, CS, MM and WTA contributed to the interpretation of the results. All authors discussed the results and contributed to the final manuscript. MM and WTA wrote the manuscript.

## Additional Information

Supplementary Information is available for this paper.

Correspondence and requests for materials should be addressed to Wesley Tyler Abplanalp.

## Data availability

The bulk RNA-seq, scRNA-seq and TCR-seq data supporting the findings of this study will be made publicly available upon acceptance of this manuscript. Source data are provided with this paper.

---

**Extended Data Figure 1:**
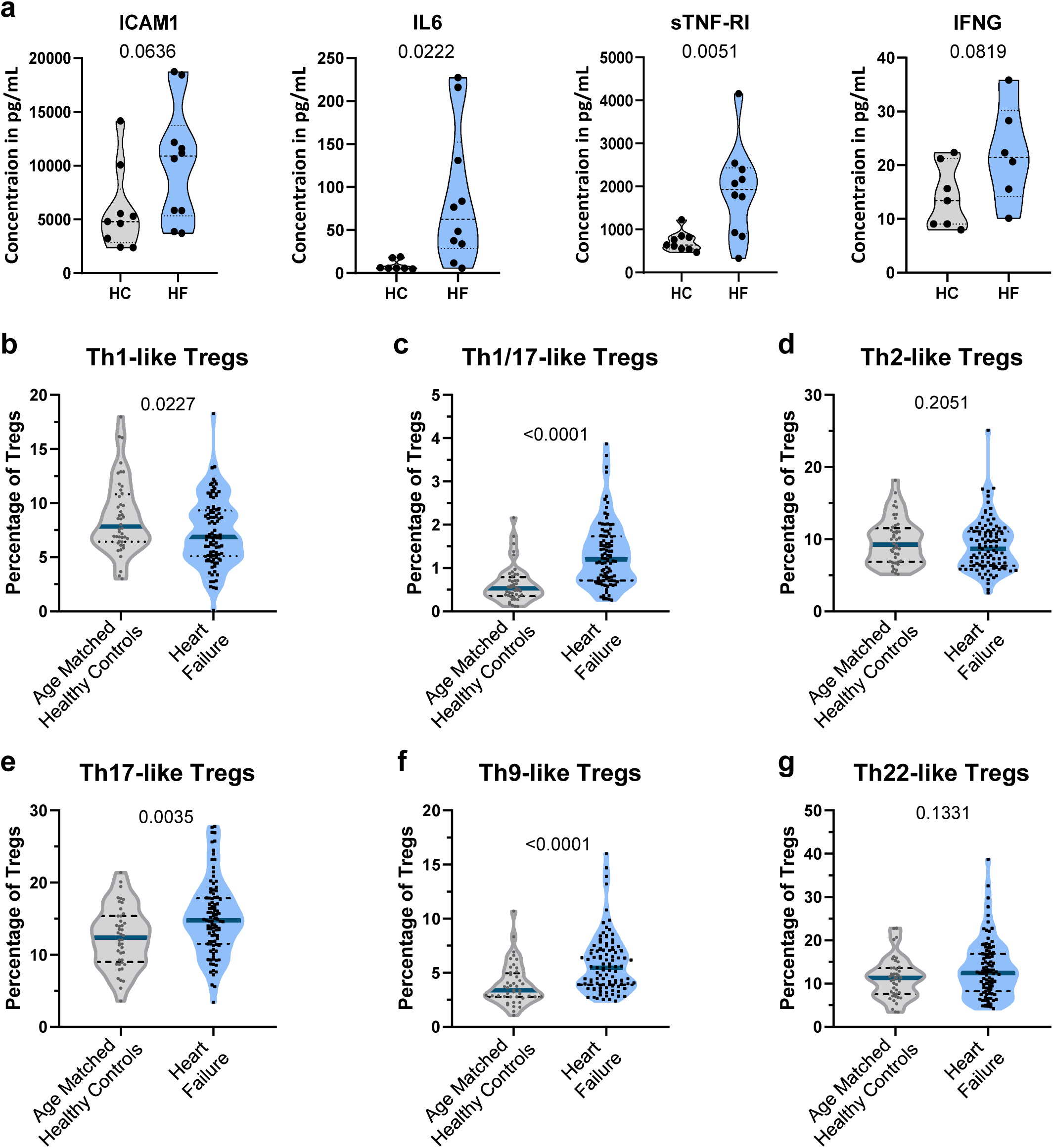
Chronic inflammation and phenotypical change of the Treg population in HF. a) Serum measurements of inflammatory markers from HF patients and HCs (Biolegend-Legendplex assay N=10 HF and N=9 HC samples). b-g) Flow cytometric deep profiling of the CD4+ Treg population within HF patients (N= 99) and age matched HCs (N=44), shown as violin plots with median and quartiles. P-values are indicated within the graphs and calculated via unpaired two sided Student’s t-test. Th - T helper cell; Treg – T regulatory cell; ICAM1 – Intracellular adhesion molecule 1; IL6 – Interleukin 6; sTNF-RI – Soluble tumor necrosis factor receptor 1; IFNG – Interferon gamma

**Extended Data Figure 2.**
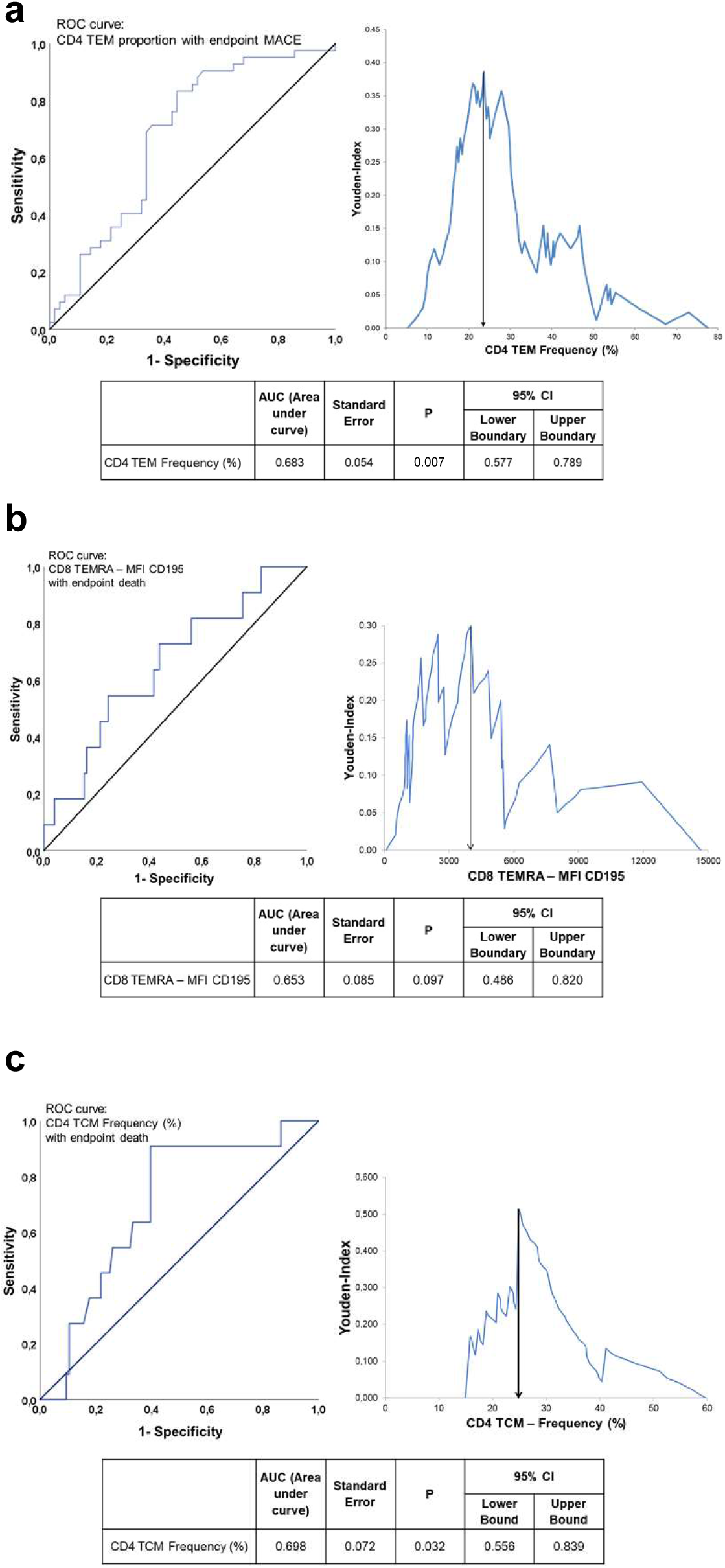
Association of immune parameters with death and MACE – correlation analysis. a) ROC curve analysis and Youden-Index to determine significant association of MACE (major adverse cardiovascular events) with CD4+ TEM cells, as well as the cut-off value for dichotomized analysis at 24% CD4+ TEM cells. b) ROC curve analysis and Youden-Index to determine association of death with the CD195 expression level on CD8+ TEMRA cells (shown as geometric mean of the MFI), as well as the cut-off value for dichotomized analysis at 4010 MFI of CD195 on CD8+ TEMRA cells. c) ROC curve analysis and Youden-Index to determine significant association of death with the CD4+ TCM frequency (shown as percent), and cut-off value for dichotomized analysis of 25% CD4+ TCM cells. Population size and group size are together with the p-values indicated within the graphs and were calculated via Mann-Whitney-U test and dichotomized Cox-regression analysis. CI – Confidence Interval; MFI – Mean fluorescence intensity; TCM – T central memory; TEM – T effector memory; TEMRA – T effector memory RA+ or terminally differentiated

**Extended Data Figure 3.**
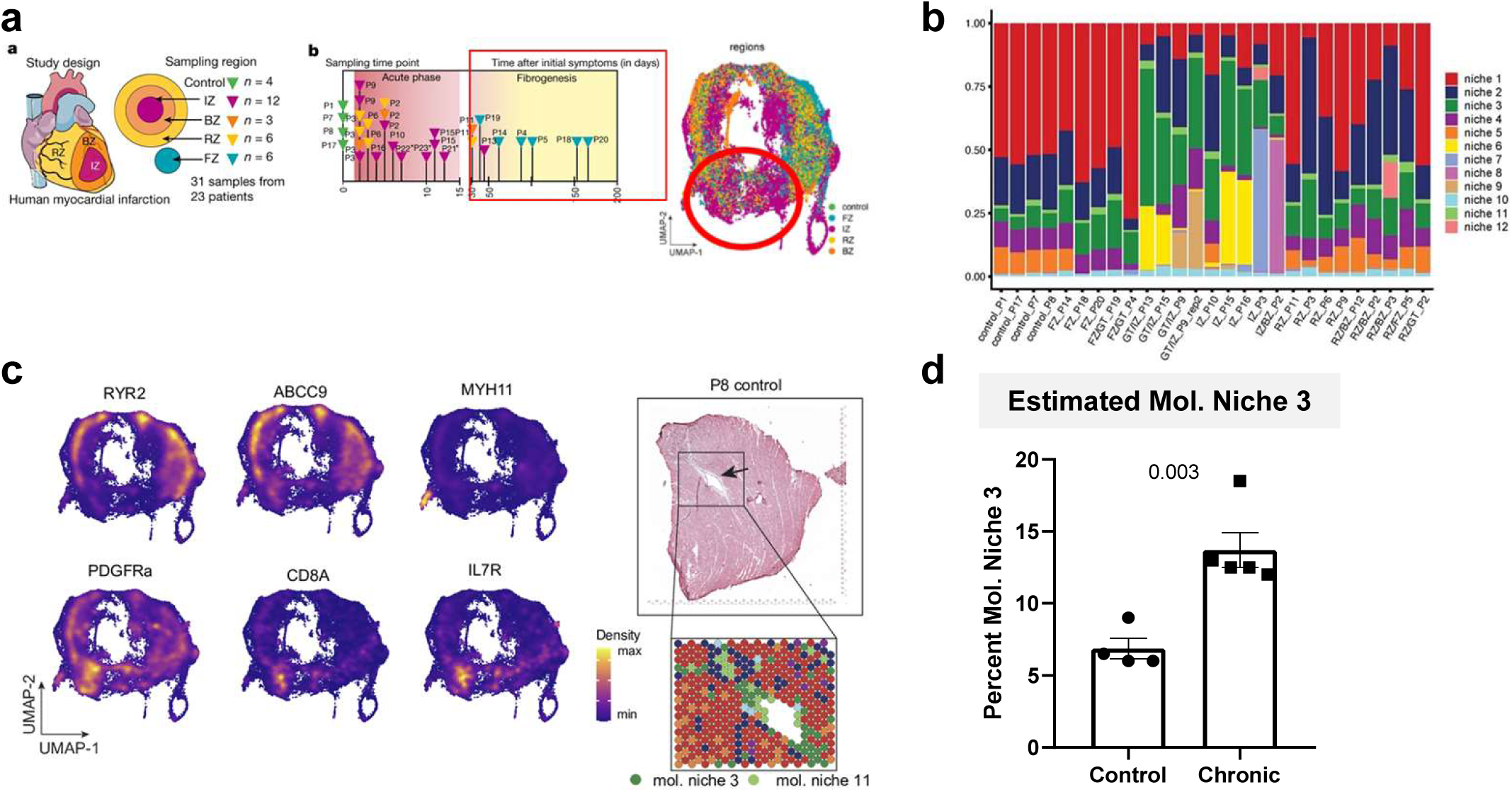
T cells infiltrate and expand within the heart. a) Overview of spatial transcriptomics analysis from human infarcted hearts at different time points. Chronic time points are highlighted with a red box (post day 30). b) Mapped and clustered molecular niches composed of different cell types, derived from all time points and sample areas post-MI, with molecular niche 3 mainly consisting of fibroblasts and immune cells. c) Location and gene expression weight (see density scale) of markers in close proximity to the infarction zone, with two of these markers being T cell associated, CD8 and IL7R. d) Relative proportion change of molecular niche 3, comparing non-infarcted hearts and chronic time points after MI (post day 30). The boxplot is shown with each donor as individual dot and the standard error of the mean (SEM). P-values are indicated within the graphs and were calculated via unpaired two sided t-test. RYR2 – Ryanodine receptor 2; ABCC9 – ATP binding cassette subfamily C member 9; MYH11 – Myosin heavy chain 11; PDGFRa - platelet derived growth factor receptor alpha; CD8A – Cluster of differentiation 8A; IL7R – CD127 = Interleukin receptor 7

**Extended Data Figure 4.**
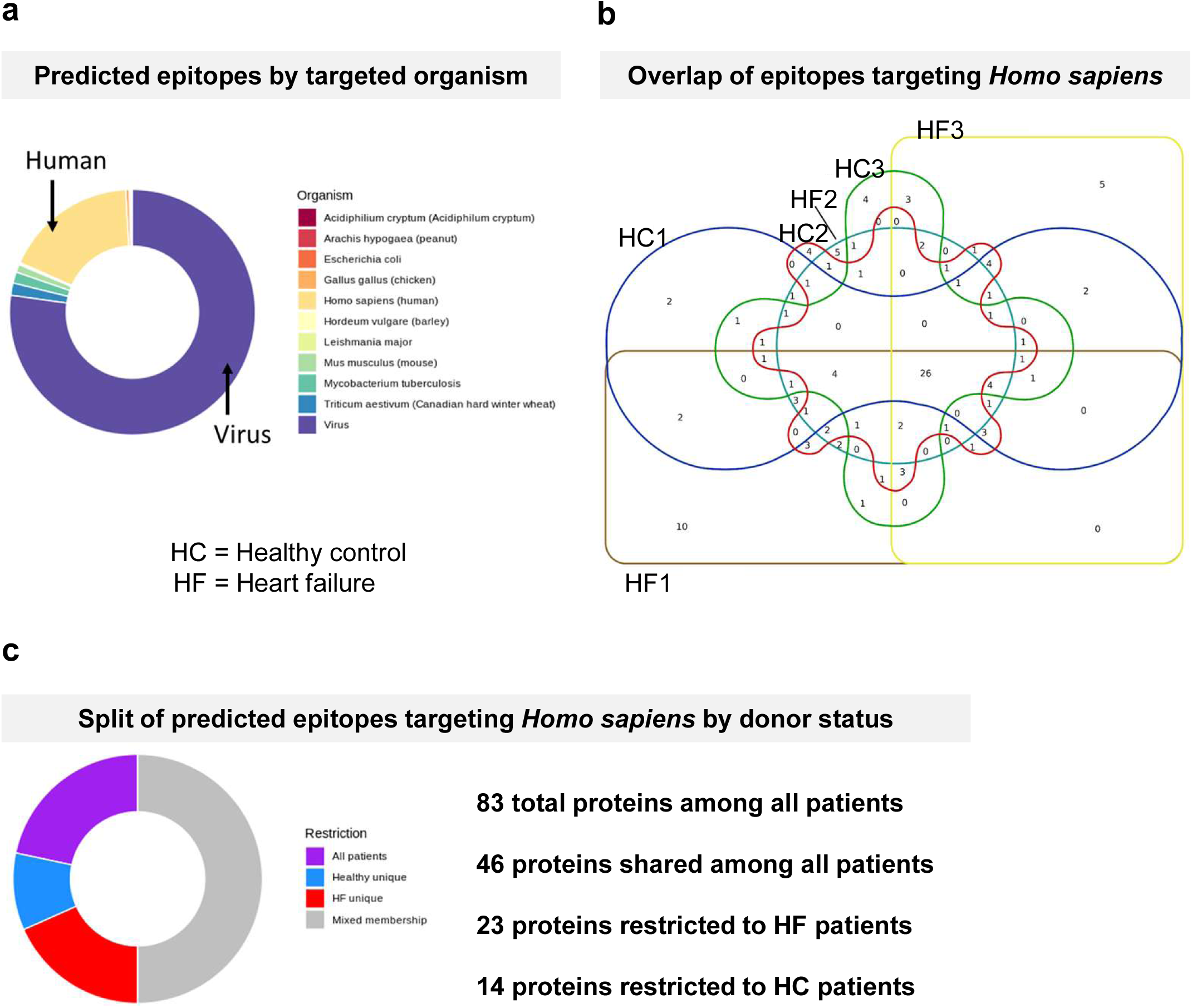
T cells of HF patients show unique predicted epitopes with reactivity to human derived antigens. a) Predicted TCR epitopes and their host origin from combined bulk TCR sequencing results, shown as the relative frequency of the antigen source proportion. Analysis was performed using TCRMatch. b) Venn diagram shows the overlap of the antigens with human origin from the different donors. c) Relative proportion of the predicted human derived antigens found within each donor (purple), only the healthy donors (blue) and only within the HF patients (red) as well as a mixed origin (grey), along with absolute number of predicted epitopes by group. The pie chart is based on the Venn diagram in b).

**Extended Data Figure 5.**
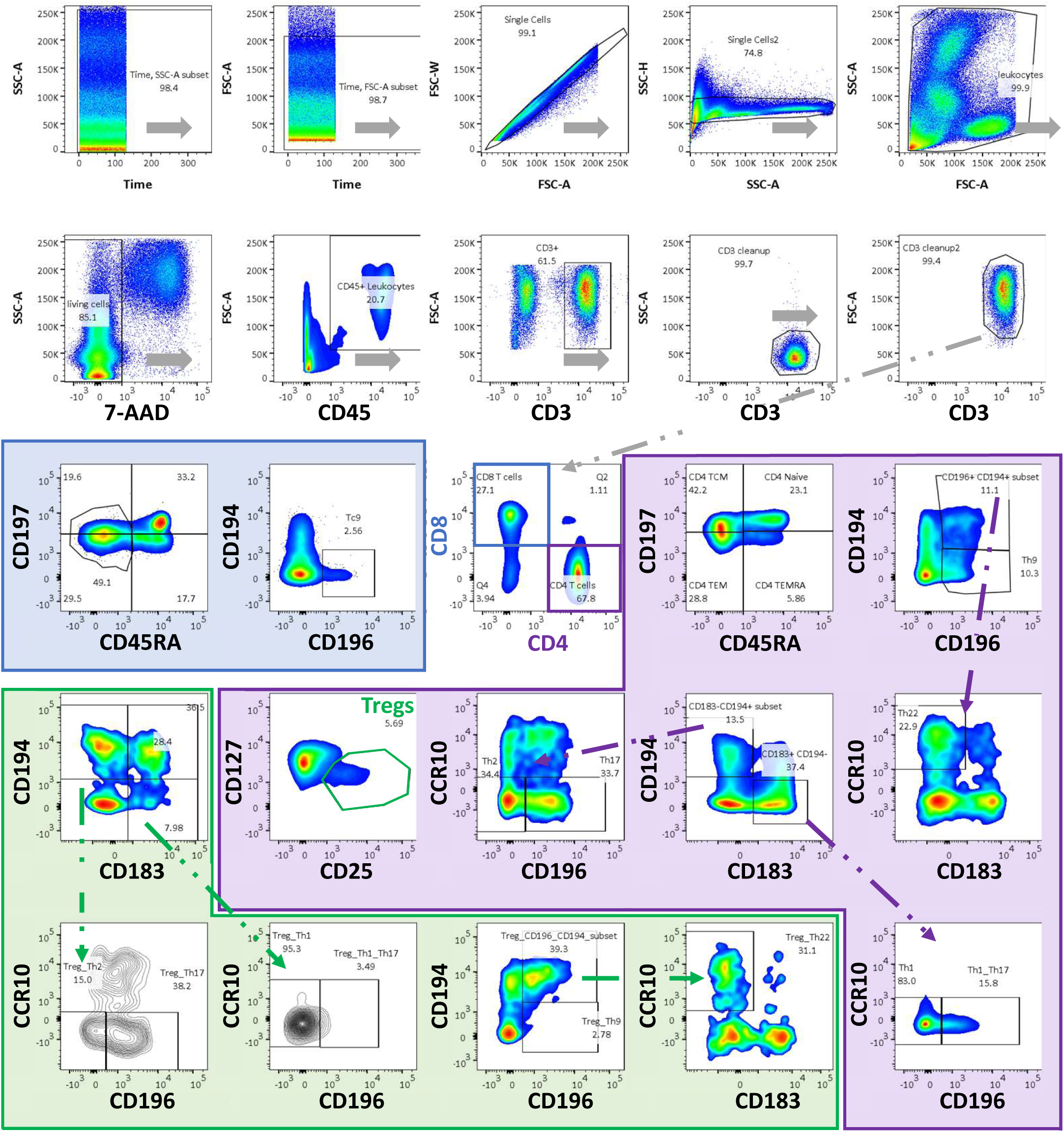
Flow cytometry gating strategy for the T cell Panel. The top two rows show the cleaning steps, including doublet removal via single cell gates (FSC-A vs. FSC-W and SSC-A vs. SSC-H). Then the target population of cells is gated (leukocytes) and living cells are selected. Verification of CD45+ expression is performed before CD3+ T cells are selected, and again cleaned. T cells are then split into CD4+ (purple box) and CD8+ (blue box). CD4+ T cells are analyzed for memory populations (purple box) via CD45RA vs. CCR7, or via CD196 vs. CD194 to obtain Th9 cells and subsequently Th22 cells (see purple arrow). CD183 vs. CD194 and the subsequent gates of CD196 vs. CCR10 are used to identify other helper subsets (see arrows). CD25 vs. CD127 shows the CD4+ Treg population (green gate). This Treg population is further analyzed, applying the same gating strategy of the CD4+ helper subsets on the Treg subpopulation (see green box, generating the Th-like Treg subpopulations). CD8+ T cells are further divided into the memory populations via CD45RA vs. CD197 and into Tc9 cells by plotting CD196 vs. CD194 (see blue box).

**Extended Data Table 1:** Patient Demographics for flow cytometry cohort. Overview HF cohort (with reduced left ventricular ejection fraction) and age matched HC. NTproBNP – N-terminal prohormone of brain natriuretic peptide

| Clinical Parameters | Patient count per parameter | HFrEF Cohort (N = 144)<br>Mean | Standard Deviation | Aged Healthy Controls (N = 44)<br>Mean | Standard Deviation |
| --- | --- | --- | --- | --- | --- |
| Age (years) | 144 | 67.5 | 10.6 | 65.0 | 8.2 |
| Body mass index | 98 | 28.3 | 5.6 | / | / |
| Blood Pressure Systolic (mmHg) | 107 | 117.7 | 19.1 | / | / |
| Left Ventricular Ejection Fraction (%) | 144 | 28.2 | 9.6 | / | / |
| New York health association score (NYHA) | 106 | 2.3 | 0.8 | / | / |
| Triglycerides (mg/dl) | 134 | 137.7 | 70.0 | / | / |
| Estimated Glomerular Filtration Rate (MDRD) | 144 | 53.7 | 21.7 | / | / |
| Creatinine (mg/dl) | 144 | 1.5 | 0.6 | / | / |
| Urea (mg/dl) | 135 | 58.7 | 32.7 | / | / |
| Albumin (g/dl) | 107 | 4.2 | 0.6 | / | / |
| Bilirubin (mg/dl) | 118 | 0.6 | 0.4 | / | / |
| NTproBNP (pg/ml) | 134 | 3135.9 | 5231.0 | / | / |
| Cholesterol (mg/dl) | 141 | 146.5 | 42.0 | / | / |
| High Density Lipoprotein -HDL (mg/dl) | 141 | 47.6 | 14.8 | / | / |
| Low Density Lipoprotein -LDL (mg/dl) | 141 | 82.2 | 35.2 | / | / |
| Creatinine kinase in serum (U/l) | 143 | 1.5 | 0.6 | / | / |
| Troponin T (pg/ml) | 120 | 62.7 | 192.4 | / | / |
| Haemoglobin (g/dl) | 144 | 13.4 | 2.0 | / | / |
| C-Reactive Protein -CRP (mg/dl) | 138 | 0.8 | 1.6 | / | / |
| Leukocytes (count/nl) | 144 | 7.9 | 1.8 | / | / |
| Thrombocytes (count/nl) | 144 | 225.7 | 76.6 | / | / |
| Follow-up time (days) | 107 | 453.8 | 242.0 | / | / |

**Extended Data Table 2:** Patient comorbidities and sex split of the flow cytometry cohort. Overview of the sex split and the tracked comorbidities of the assessed HF cohort (with reduced left ventricular ejection fraction) and age matched HC. NTproBNP – N-terminal prohormone of brain natriuretic peptide

| Clinical Characteristics and Comorbidities | Patient count per parameter | HFrEF Cohort (N = 144) | % from Total | Aged Healthy Controls (N = 44) | % from Total |
| --- | --- | --- | --- | --- | --- |
| **Sex** | **144** |  |  |  |  |
| Female |  | 21 | 14.6 | 22 | 50.0 |
| Male |  | 123 | 85.4 | 22 | 50.0 |
| **Number of diseased vessels** | **144** |  |  |  |  |
| 0 |  | 1 | 0.7 | / | / |
| 1 |  | 33 | 22.9 | / | / |
| 2 |  | 38 | 26.4 | / | / |
| 3 |  | 72 | 50.0 | / | / |
| **History of Myocardial Infarction** | **144** |  |  |  |  |
| 0 |  | 48 | 33.3 | / | / |
| 1-2 |  | 96 | 66.7 | / | / |
| History of Percutaneous Coronary Intervention (PCI) | **143** | 117 | 81.3 | / | / |
| History of Coronary Artery Bypass Graft Surgery (CABG) | **144** | 46 | 31.9 | / | / |
| Implantable Cardioverter-Defibrillator (ICD) | **144** | 73 | 50.7 | / | / |
| Cardiac Resynchronization Therapy (CRT) | **144** | 28 | 19.4 | / | / |
| Diabetes Mellitus | **142** | 53 | 36.8 | / | / |
| Hyperlipidemia | **116** | 78 | 54.2 | / | / |
| **Smoker** | **144** |  |  |  |  |
| Never |  | 70 | 48.6 | / | / |
| Actively |  | 41 | 28.5 | / | / |
| Stopped/Ex-Smoker |  | 33 | 22.9 | / | / |
| **Adiposity** | **143** | 42 | 29.2 | / | / |
| Hypertension | **144** | 109 | 75.7 | / | / |
| Hyperuricemia | **116** | 41 | 28.5 | / | / |
| Renal Failure (GFR <60 ml/min) | **144** | 80 | 55.6 | / | / |
| Peripheral Arterial Disease (PAD) | **143** | 30 | 20.8 | / | / |
| Stroke/Transient Ischemic Attack (TIA) | **144** | 19 | 13.2 | / | / |

**Extended Data Table 3.**
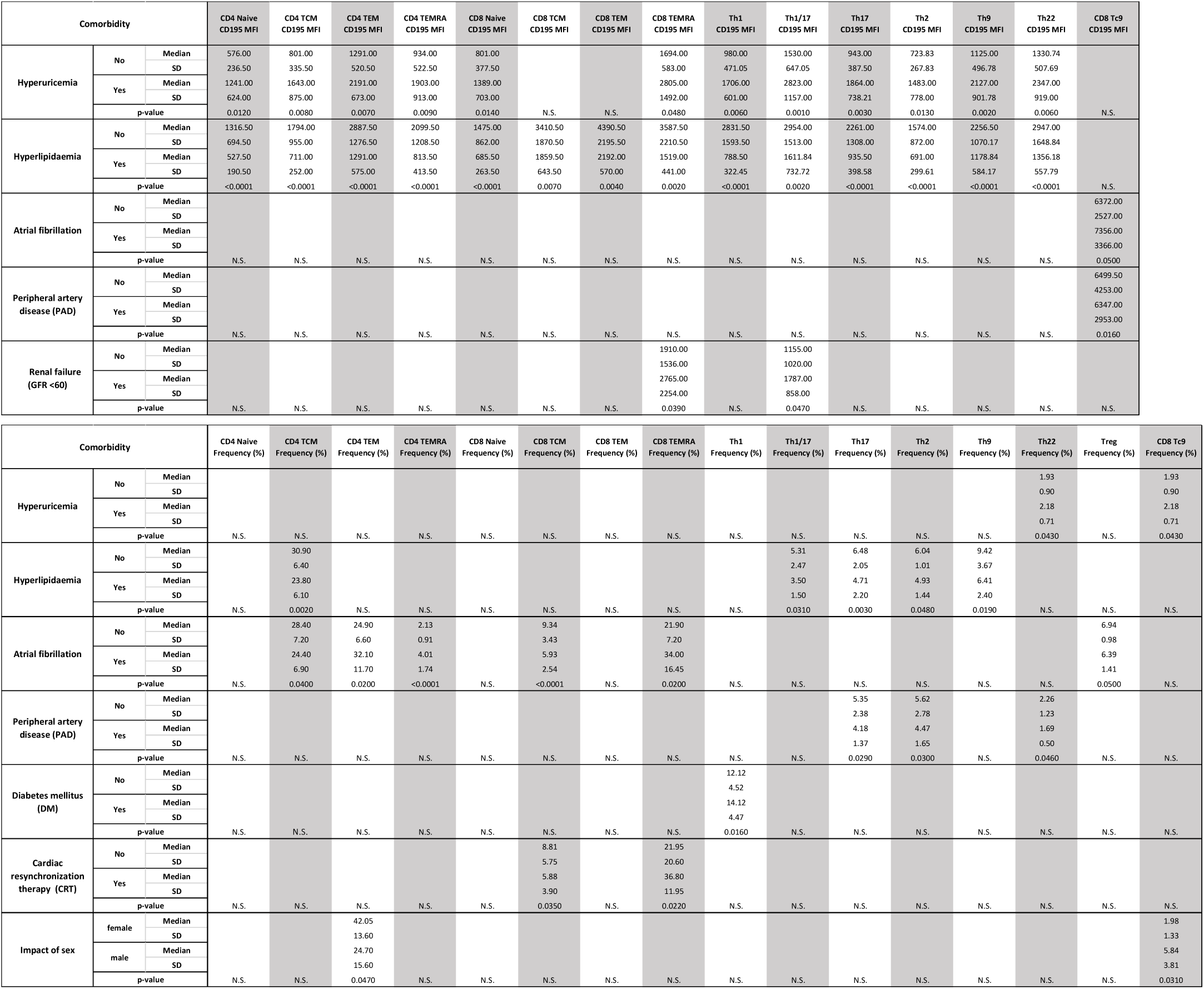
Association of clinical comorbidities with immune composition changes. Overview of the multivariate regression analysis considering clinical comorbidities that do significantly associate with the changes of the T cell compartment within the HF cohort. Only significant associations between the comorbidity and certain T cell subset frequencies or their mean CCR5 expression levels (MFI) are shown. CD4 and CD8 subset frequencies are always referred to their respective CD4 or CD8 parent population. SD – Standard deviation; N.S. – Not significant p-values; GFR – Glomerular Filtration Rate

**Extended Data Table 4:**
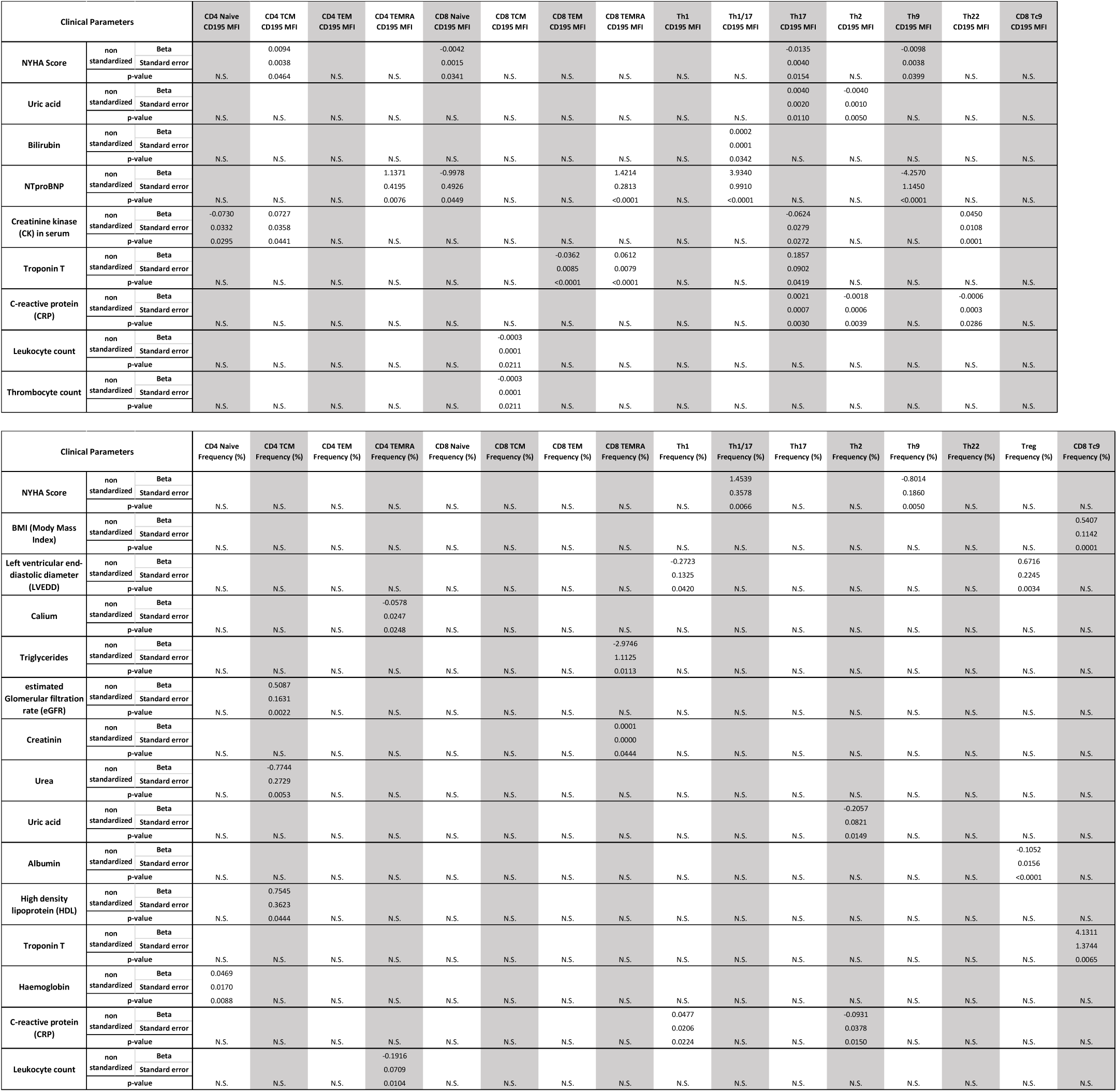
Association of assessed clinical parameters with immune composition changes. Overview of the multivariate regression analysis considering clinical blood/serum parameters that do significantly associate with changes of the T cell compartment within the HF cohort. Only significant associations between assessed clinical parameters and certain T cell subset frequencies or their mean CCR5 expression levels (MFI) are shown. Beta gives the slope of the correlation and thus the effect size. CD4 and CD8 subset frequencies are always referred to their respective CD4 or CD8 parent population. N.S. – Not significant p-values; NYHA – New York Health Association

**Extended Data Table 5:** Associations of immune and clinical parameters with patients death. Overview of the associations between death and the assessed clinical and immunological parameters within the HF cohort. HF patients were split into survivors and dead patients, comparing the two groups for each parameter/T cell subset individually using an unpaired two-sided t-test. Significant associations are highlighted with bold letters. CD4 and CD8 subset frequencies are always referred to their respective CD4 or CD8 parent population. NTproBNP – N-terminal prohormone of brain natriuretic peptide; CRP – C-reactive protein; TCM – T central memory cell; TEM – T effector memory cell; TEMRA – T effector memory CD45RA+ cell; Treg – T regulatory cell; Tc9 – T cytotoxic cell 9

| HFREF cohort with information about death (N=107) |  |  |  |  |
| --- | --- | --- | --- | --- |
| Immune and clinical parameters<br>(Mean ± SD) | Patient count per<br>Parameter | Death (N=11) | Survivors (N=96) | p-value |
| NTproBNP pg/ml | 100 | 7199 ± 9132.5 | 1068.7 ± 3899 | 0.0000 |
| Troponin T pg/ml | 88 | 68.7 ± 310.4 | 24.8 ± 37.5 | 0.0002 |
| Hemoglobin g/dl | 107 | 10.6 ± 1.2 | 13.5 ± 1.9 | 0.0000 |
| CRP mg/dl | 101 | 0.9 ± 3.2 | 0.3 ± 1 | 0.0003 |
| Leukocyte count/nl | 107 | 6.6 ± 1.3 | 7.6 ± 1.9 | 0.0575 |
| Estimated Glomerular Filtration Rate (MDRD) | 107 | 30.8 ± 20.8 | 51.7 ± 22.7 | 0.0041 |
| Creatinine mg/dl | 107 | 2.1 ± 0.8 | 1.3 ± 0.5 | 0.0000 |
| Urea mg/dl | 101 | 80.4 ± 47.3 | 48.4 ± 28.8 | 0.0002 |
| Uric Acid mg/dl | 56 | 10.8 ± 2.3 | 6.6 ± 2.3 | 0.0001 |
| Albumin g/dl | 80 | 4 ± 0.3 | 4.2 ± 0.4 | 0.1340 |
| Cholesterol mg/dl | 107 | 126.9 ± 31.4 | 144.8 ± 41.7 | 0.1427 |
| Body Mass Index (BMI) | 74 | 29.7 ± 11.7 | 28 ± 4.8 | 0.1978 |
| CD4 Naive Frequency (%) | 107 | 41.1 ± 21.8 | 33.9 ± 15.7 | 0.1032 |
| CD4 TCM Frequency (%) | 107 | 20.6 ± 7.3 | 25.7 ± 12.1 | 0.0636 |
| CD4 Effector/Memory Frequency (%) | 107 | 48.5 ± 21.8 | 59.9 ± 15.7 | 0.1026 |
| Treg Frequency (%) | 107 | 4.9 ± 2.8 | 7 ± 3.2 | 0.0738 |
| CD4 TEMRA CD195 MFI | 106 | 1603.1 ± 1953.3 | 1271.2 ± 1591.7 | 0.1857 |
| CD8 TEM CD195 MFI | 107 | 4570.6 ± 2410.4 | 2838.7 ± 2650.3 | 0.0703 |
| **CD8 TEMRA CD195 MFI** | **107** | **3507.3 ± 3745.5** | **2117.2 ± 2111.7** | **0.0145** |
| **CD8 Tc9 CD195 MFI** | **106** | **10925.2 ± 7375.5** | **5618.7 ± 6406.4** | **0.0268** |

**Extended Data Table 6:** Associations of immune and clinical parameters with MACE. Overview of the associations between major cardiac adverse vents (MACE) and the assessed clinical and immunological parameters within the HF cohort. HF patients were split into patients with MACE and without MACE (no MACE), comparing the two groups for each parameter/T cell subset individually using an unpaired two-sided t-test. Significant associations are highlighted with bold letters. CD4 and CD8 subset frequencies are always referred to their respective CD4 or CD8 parent population. NTproBNP – N-terminal prohormone of brain natriuretic peptide; TCM – T central memory cell; TEM – T effector memory cell; TEMRA – T effector memory CD45RA+ cell; Treg – T regulatory cell; Th17 – T helper cell 17; MFI – Mean fluorescence intensity

| HFrEF cohort with information about MACE (N=113) |  |  |  |  |
| --- | --- | --- | --- | --- |
| Immune and clinical parameters<br>(Mean ± SD) | Patient count per<br>Parameter | MACE (N=50) | no MACE (N=63) | p-value |
| NTproBNP pg/ml | 106 | 1935.6 ± 5737.4 | 934.4 ± 4335.5 | 0.0785 |
| Troponin T pg/ml | 94 | 32.4 ± 159.7 | 24.9 ± 29.7 | 0.1116 |
| Thrombocyte counts/nl | 113 | 223.6 ± 82.2 | 206.2 ± 70 | 0.1718 |
| Estimated Glomerular Filtration Rate (GFR) | 113 | 45.6 ± 21 | 51.4 ± 22.6 | 0.1846 |
| Creatinine mg/dl | 113 | 1.5 ± 0.7 | 1.3 ± 0.5 | 0.1491 |
| Urea mg/dl | 107 | 54.2 ± 36.9 | 48.7 ± 28.3 | 0.1915 |
| Uric Acid mg/dl | 58 | 7.6 ± 3.1 | 6.6 ± 2.2 | 0.0709 |
| CD4 Naive Frequency (%) | 113 | 31.1 ± 16.5 | 35.6 ± 16.7 | 0.1544 |
| CD4 TCM Frequency (%) | 113 | 24.6 ± 11.4 | 26.5 ± 12.7 | 0.1809 |
| **CD4 TEM Frequency (%)** | **113** | **28.9 ± 15.2** | **22.3 ± 13.7** | **0.0053** |
| TH17 Frequency (%) | 113 | 4.8 ± 3 | 4.1 ± 2.8 | 0.1445 |
| CD8 TEMRA CD195 MFI | 113 | 2595.8 ± 2877 | 2153.7 ± 2064.9 | 0.1548 |

**Extended Data Table 7:** Association of immune parameters with death – univariate Cox regression analysis. Overview of the associations between death and the assessed clinical and immunological parameters within the HF cohort in a Cox regression model, displaying the hazard ratio with an the confidential Interval (CI). CD4 and CD8 subset frequencies are always referred to their respective CD4 or CD8 parent population. * - HR is related to 1000 units of the MFI or the logarithmic NTproBNP levels; ** - Gives the number of patients where the investigators had information about the given parameter e.g. presence of absence of a disease/assist device; *** - Dichotomized Hazard ratio and p-values: NTproBNP – N-terminal prohormone of brain natriuretic peptide; TCM – T central memory cell; TEM – T effector memory cell; TEMRA – T effector memory CD45RA+ cell; Th – T helper cell; Tc – T cytotoxic cell; MFI – Mean fluorescence intensity; df – degree of freedom

| Parameter assessed in correlation with patient death | patient count per parameter** | Beta | Standard Error | Wald | df | p-value | Hazard Ratio (HR) | 95.0% CI for HR |  |
| --- | --- | --- | --- | --- | --- | --- | --- | --- | --- |
|  |  |  |  |  |  |  |  | Lower Boundary | Upper Boundary |
| NTproBNP - pg/ml | 100 | 0.0001 | 0.0000 | 19.4754 | 1.0 | 0.0000 | 1.0001 | 1.0001 | 1.0002 |
| Hemoglobin - g/dl | 107 | -0.7051 | 0.1633 | 18.6404 | 1.0 | 0.0000 | 0.4941 | 0.3587 | 0.6804 |
| NTproBNP (Log)* | 100 | 2.9165 | 0.7294 | 15.9858 | 1.0 | 0.0001 | 18.4761 | 4.4229 | 77.1818 |
| Urea - mg/dl | 101 | 0.0241 | 0.0069 | 12.1860 | 1.0 | 0.0005 | 1.0244 | 1.0106 | 1.0384 |
| Troponin T - pg/ml | 88 | 0.0050 | 0.0014 | 11.8163 | 1.0 | 0.0006 | 1.0050 | 1.0021 | 1.0078 |
| Uric Acid - mg/dl | 56 | 0.5432 | 0.1586 | 11.7313 | 1.0 | 0.0006 | 1.7215 | 1.2616 | 2.3491 |
| New York Health Association Score (NYHA) | 99 | 1.5020 | 0.5256 | 8.1675 | 1.0 | 0.0043 | 4.4906 | 1.6031 | 12.5792 |
| Estimated Glomerular Filtration Rate (MDRD) | 107 | -0.0577 | 0.0211 | 7.4674 | 1.0 | 0.0063 | 0.9439 | 0.9057 | 0.9838 |
| CD8 TEMRA CD195 MFI >4010 *** | 109 | 0.2673 | 0.1049 | 6.4980 | 1.0 | 0.0353 | 3.6028 | 1.0924 | 11.8824 |
| Chronic Obstructive Pulmonary Disease (COPD) | 107 | 1.6581 | 0.6569 | 6.3716 | 1.0 | 0.0116 | 5.2494 | 1.4487 | 19.0215 |
| History of Percutaneous Coronary Intervention (PCI) | 107 | -1.3745 | 0.6060 | 5.1439 | 1.0 | 0.0233 | 0.2530 | 0.0771 | 0.8297 |
| CD4 TCM - Frequency <25% *** | 107 | -0.0606 | 0.0292 | 4.3037 | 1.0 | 0.0084 | 16.0200 | 2.0386 | 125.8903 |
| Renal Failure (GFR <60 ml/min) | 107 | 1.9794 | 1.0496 | 3.5567 | 1.0 | 0.0593 | 7.2385 | 0.9252 | 56.6297 |
| Leukocyte - count/nl blood | 107 | -0.3190 | 0.1780 | 3.2110 | 1.0 | 0.0731 | 0.7268 | 0.5127 | 1.0304 |
| BMI - Body Mass Index | 74 | 0.1049 | 0.0618 | 2.8822 | 1.0 | 0.0896 | 1.1105 | 0.9839 | 1.2535 |
| Albumin - g/dl | 80 | -0.8719 | 0.5298 | 2.7083 | 1.0 | 0.0998 | 0.4182 | 0.1480 | 1.1812 |
| CD8 TEM CD195 MFI* | 107 | 0.1490 | 0.0924 | 2.6012 | 1.0 | 0.1068 | 1.1607 | 0.9684 | 1.3912 |
| CD8 Tc9 CD195 MFI* | 106 | 0.0543 | 0.0337 | 2.5965 | 1.0 | 0.1071 | 1.0558 | 0.9883 | 1.1280 |
| Cholesterol - mg/dl | 107 | -0.0536 | 0.0345 | 2.4216 | 1.0 | 0.1197 | 0.9478 | 0.8859 | 1.0140 |
| Potassium - mmol/l | 108 | -0.9424 | 0.6197 | 2.3126 | 1.0 | 0.1283 | 0.3897 | 0.1157 | 1.3129 |
| Atrial fibrillation | 107 | 0.9282 | 0.6279 | 2.1856 | 1.0 | 0.1393 | 2.5300 | 0.7391 | 8.6610 |
| Hyperlipidemia | 88 | -0.9350 | 0.6737 | 1.9260 | 1.0 | 0.1652 | 0.3926 | 0.1048 | 1.4703 |
| Peripheral Artery Disease | 108 | 0.8519 | 0.6456 | 1.7410 | 1.0 | 0.1870 | 2.3440 | 0.6613 | 8.3084 |
| Low Density Lipoprotein - mg/dl | 107 | 0.0434 | 0.0364 | 1.4186 | 1.0 | 0.2336 | 1.0443 | 0.9724 | 1.1215 |
| Hyperuricemia | 87 | 0.7452 | 0.6600 | 1.2747 | 1.0 | 0.2589 | 2.1068 | 0.5779 | 7.6812 |
| Creatinine Kinase (CK) Serum - U/l | 106 | -0.0070 | 0.0070 | 1.0081 | 1.0 | 0.3154 | 0.9930 | 0.9794 | 1.0067 |
| Left Ventricular Enddiastolic Diameter - mm | 105 | 0.0393 | 0.0392 | 1.0004 | 1.0 | 0.3172 | 1.0400 | 0.9630 | 1.1232 |
| High Density Lipoprotein - mg/dl | 107 | 0.0253 | 0.0317 | 0.6350 | 1.0 | 0.4255 | 1.0256 | 0.9638 | 1.0913 |
| History of MI | 107 | -0.3186 | 0.4241 | 0.5646 | 1.0 | 0.4524 | 0.7271 | 0.3167 | 1.6695 |
| Coronary Artery Disease (CAD) | 96 | 0.2953 | 0.3959 | 0.5564 | 1.0 | 0.4557 | 1.3436 | 0.6184 | 2.9193 |
| Bypass | 107 | 0.4495 | 0.6058 | 0.5506 | 1.0 | 0.4581 | 1.5675 | 0.4782 | 5.1387 |
| Arterial Hypertension | 97 | -0.5064 | 0.6905 | 0.5379 | 1.0 | 0.4633 | 0.6026 | 0.1557 | 2.3324 |
| Triglycerides | 102 | -0.0037 | 0.0056 | 0.4365 | 1.0 | 0.5088 | 0.9963 | 0.9855 | 1.0073 |
| Diabetes Mellitus | 106 | 0.3306 | 0.6076 | 0.2960 | 1.0 | 0.5864 | 1.3918 | 0.4230 | 4.5792 |
| Stroke/Transient Ischemic Attack (TIA) | 107 | 0.2425 | 0.7820 | 0.0962 | 1.0 | 0.7565 | 1.2744 | 0.2752 | 5.9014 |
| Adiposity | 85 | 0.2027 | 0.7644 | 0.0703 | 1.0 | 0.7909 | 1.2247 | 0.2737 | 5.4791 |
| Sodium - mmol/l | 106 | 0.0048 | 0.0295 | 0.0265 | 1.0 | 0.8707 | 1.0048 | 0.9484 | 1.0646 |
| Systolic Blood Pressure - mmHg | 82 | 0.0019 | 0.0150 | 0.0161 | 1.0 | 0.8989 | 1.0019 | 0.9729 | 1.0318 |
| Left Ventricular Ejection Fraction - % | 107 | 0.0039 | 0.0331 | 0.0141 | 1.0 | 0.9054 | 1.0039 | 0.9408 | 1.0713 |
| Thrombocyte - count/nl blood | 107 | 0.0004 | 0.0038 | 0.0106 | 1.0 | 0.9180 | 1.0004 | 0.9930 | 1.0079 |
| Bilirubin - mg/dl | 89 | 0.0965 | 0.9469 | 0.0104 | 1.0 | 0.9188 | 1.1013 | 0.1721 | 7.0459 |

**Extended Data Table 8:** Association of immune parameters with MACE – binary logistic regression and multivariate analysis. Overview of the associations between MACE and the assessed clinical and immunological parameters within the HF cohort in binary logistic regression analysis, displaying the odds ratio and the confidential Interval (CI). CD4 and CD8 subset frequencies are always referred to their respective CD4 or CD8 parent population. * - HR is related to 1000 units of the MFI or the logarithmic NTproBNP levels; ** - Gives the number of patients where the investigators had information about the given parameter e.g. presence of absence of a disease/assist device; *** - Dichotomized Hazard ratio and p-values; NTproBNP – N-terminal prohormone of brain natriuretic peptide; TCM – T central memory cell; TEM – T effector memory cell; TEMRA – T effector memory CD45RA+ cell; Treg – T regulatory cell; Th – T helper cell; TC – T cytotoxic cell; MFI – Mean fluorescence intensity

| HFREF cohort MACE binary logistic regression analysis (N=113) |  | Univariate |  |  | Multivariate |  |  |
| --- | --- | --- | --- | --- | --- | --- | --- |
| Immune and clinical parameters | Patient count per Parameter** | p-value | Odds Ratio | 95% Confidence Interval | p-value | Odds Ratio | 95% Confidence Interval |
| NTproBNP - Log * | 106 | 0.0051 | 2.9525 | 1.3841 - 6.2984 | 0.0006 | 5.3833 | 2.0687 - 14.0091 |
| CD4 TEM Frequency >24% *** | 113 | 0.0054 | 3.1909 | 1.4095 - 7.2236 | 0.0018 | 5.0091 | 1.8214 - 13.776 |
| Hemoglobin - g/dl | 113 | 0.0099 | 0.7690 | 0.6299 - 0.9388 | 0.3542 | 0.8851 | 0.6836 - 1.1459 |
| Chronic Obstructive Pulmonary Disease | 113 | 0.0122 | 4.6579 | 1.399 - 15.5079 | 0.0622 | 3.6385 | 0.9362 - 14.1412 |
| New York Health Association Score | 84 | 0.0277 | 1.8455 | 1.0695 - 3.1848 | 0.2481 | 1.4529 | 0.7707 - 2.7387 |
| Low Density Lipoprotein - mg/dl | 113 | 0.1418 | 1.0314 | 0.9897 - 1.0747 |  |  |  |
| CD8 Naive CD195 MFI* | 113 | 0.1614 | 0.9989 | 0.9974 - 1.0004 |  |  |  |
| Cardiac Resynchronization Therapy | 113 | 0.1664 | 0.5154 | 0.2016 - 1.3177 |  |  |  |
| Atrial Fibrillation | 113 | 0.1750 | 1.7484 | 0.7798 - 3.9202 |  |  |  |
| Thrombocyte - count/nl blood | 113 | 0.1779 | 1.0035 | 0.9984 - 1.0086 |  |  |  |
| CD4 TCM Frequency - % | 113 | 0.1813 | 0.9788 | 0.9486 - 1.01 |  |  |  |
| Cholesterol - mg/dl | 113 | 0.2060 | 0.9753 | 0.9383 - 1.0138 |  |  |  |
| estimated Glomerular Filtration Rate (MDRD) | 113 | 0.2400 | 0.9892 | 0.9714 - 1.0073 |  |  |  |
| Th17 Frequency - % | 113 | 0.2741 | 1.1667 | 0.8851 - 1.5379 |  |  |  |
| Leukocyte - count/nl blood | 113 | 0.2753 | 0.8903 | 0.7225 - 1.097 |  |  |  |
| Th1 CD195 MFI* | 109 | 0.3392 | 1.0003 | 0.9997 - 1.0009 |  |  |  |
| CD4 TEMRA CD195 MFI* | 112 | 0.3575 | 1.0003 | 0.9996 - 1.001 |  |  |  |
| Left Bundle Branch Block | 113 | 0.3612 | 0.6470 | 0.2541 - 1.6474 |  |  |  |
| CD8 TEMRA CD195 MFI* | 113 | 0.3789 | 1.1883 | 0.8092 - 1.745 |  |  |  |
| Renal Failure (GFR<60 ml/min) | 113 | 0.3838 | 1.4188 | 0.6457 - 3.1173 |  |  |  |
| CD4 TCM CD195 MFI* | 113 | 0.3894 | 1.0012 | 0.9984 - 1.0041 |  |  |  |
| CD8 TCM CD195 MFI* | 113 | 0.3998 | 0.9998 | 0.9993 - 1.0003 |  |  |  |
| High Density Lipoprotein - mg/dl | 113 | 0.4073 | 1.0155 | 0.9792 - 1.0532 |  |  |  |
| Th1/17 CD195 MFI* | 110 | 0.4186 | 1.0004 | 0.9994 - 1.0015 |  |  |  |
| History of Myocardial Infarction | 113 | 0.4552 | 1.2396 | 0.7054 - 2.1783 |  |  |  |
| Right Bundle Branch Block | 113 | 0.4692 | 1.7057 | 0.4017 - 7.2426 |  |  |  |
| CD8 TEMRA Frequency - % | 113 | 0.4775 | 1.0073 | 0.9873 - 1.0278 |  |  |  |
| Th17 CD195 MFI* | 110 | 0.5038 | 0.9993 | 0.9971 - 1.0014 |  |  |  |
| Implantable Cardioverter Defibrillator | 113 | 0.5109 | 0.7702 | 0.3537 - 1.6774 |  |  |  |
| C-Reactive Protein - mg/dl | 107 | 0.5178 | 1.0894 | 0.8404 - 1.4122 |  |  |  |
| Th22 Frequency - % | 113 | 0.5409 | 0.8838 | 0.5947 - 1.3133 |  |  |  |
| Treg Frequency - % | 113 | 0.5605 | 1.0385 | 0.9145 - 1.1792 |  |  |  |
| Th1 Frequency - % | 113 | 0.5876 | 1.0211 | 0.9469 - 1.101 |  |  |  |
| CD4 Naive CD195 MFI* | 112 | 0.6150 | 0.9993 | 0.9966 - 1.002 |  |  |  |
| Tc9 CD195 MFI* | 112 | 0.6335 | 1.0000 | 0.9999 - 1.0001 |  |  |  |
| History of Percutaneous Coronary Interventions | 113 | 0.6600 | 0.7935 | 0.2832 - 2.2236 |  |  |  |
| Th9 CD195 MFI* | 110 | 0.6616 | 0.9997 | 0.9985 - 1.001 |  |  |  |
| Stroke/Transient Ischemic Attack (TIA) | 113 | 0.6796 | 1.2704 | 0.408 - 3.9556 |  |  |  |
| Diabetes Mellitus | 113 | 0.6826 | 1.1864 | 0.523 - 2.6914 |  |  |  |
| CD4 TEMRA Frequency - % | 113 | 0.6954 | 0.9853 | 0.915 - 1.061 |  |  |  |
| Left Ventricular Ejection Fraction - % | 113 | 0.7747 | 0.9934 | 0.9494 - 1.0395 |  |  |  |
| CD8 TCM Frequency - % | 113 | 0.7914 | 2.8631 | 0.0012 - 6956.0199 |  |  |  |
| CD8 TEM Frequency - % | 113 | 0.7916 | 2.8586 | 0.0012 - 6914.5175 |  |  |  |
| CD8 Naive Frequency - % | 113 | 0.7918 | 2.8552 | 0.0012 - 6901.3699 |  |  |  |
| Th1/17 Frequency - % | 113 | 0.7988 | 1.0191 | 0.8809 - 1.179 |  |  |  |
| History of Coronary Artery Bypass Graft Surgery (CABG) | 113 | 0.8120 | 0.9021 | 0.3862 - 2.1075 |  |  |  |
| Th2 CD195 MFI* | 110 | 0.8266 | 1.0002 | 0.9984 - 1.0019 |  |  |  |
| Th9 Frequency - % | 113 | 0.8607 | 0.9917 | 0.9032 - 1.0889 |  |  |  |
| Th22 CD195 MFI* | 110 | 0.8741 | 1.0001 | 0.9992 - 1.001 |  |  |  |
| CD4 TEM CD195 MFI* | 113 | 0.9128 | 0.9999 | 0.9988 - 1.0011 |  |  |  |
| Creatinine Kinase Serum - U/l | 112 | 0.9154 | 1.0003 | 0.9942 - 1.0064 |  |  |  |
| CD4 Naive Frequency - % | 113 | 0.9604 | 0.8237 | 0.0004 - 1752.6583 |  |  |  |
| CD4 Effector/Memory cell Frequency - % | 113 | 0.9637 | 0.8368 | 0.0004 - 1789.4306 |  |  |  |
| Th2 Frequency - % | 113 | 0.9793 | 1.0017 | 0.8819 - 1.1378 |  |  |  |
| Left Ventricular End-Diastolic Diameter - mm | 111 | 0.9949 | 1.0002 | 0.9511 - 1.0517 |  |  |  |
| CD8 TEM CD195 MFI* | 113 | 0.9968 | 0.9992 | 0.6719 - 1.4859 |  |  |  |

**Extended Data Table 9:** Antibodies used for flow cytometry measurement of peripheral blood.

| Antibody-targeted Antigen | Dilution | Clone | Fluorochrome | Customer | Catalogue Number |
| --- | --- | --- | --- | --- | --- |
| CD3 | 1:20 | OKT3 | BV786 | Biolegend | 317330 |
| CD4 | 1:20 | RPA-T4 | FITC | Biolegend | 300506 |
| CCR10 | 1:20 | 1B5 | PE | BD Biosciences | 563656 |
| CD8 | 1:20 | RPA-T8 | PerCP-Cy5.5 | Biolegend | 301032 |
| CD25 | 1:20 | BC96 | PE-dazzle | Biolegend | 302646 |
| CD45 | 1:20 | HI30 | BUV395 | BD Biosciences | 563792 |
| CD127 | 1:20 | A019D5 | BV510 | Biolegend | 351332 |
| CD183 | 1:20 | G025H7 | APC-Cy7 | Biolegend | 353722 |
| CD194 | 1:20 | L291H4 | PE-Cy7 | Biolegend | 359410 |
| CD195 | 1:20 | J418F1 | BV421 | Biolegend | 359118 |
| CD196 | 1:20 | G034E3 | AF647 | Biolegend | 353404 |
| CD197 | 1:20 | G043H7 | BV605 | Biolegend | 353224 |
| CD45RA | 1:50 | HI100 | AF700 | Biolegend | 304120 |

**Supplementary Table 1:** Association of immune parameters with MACE – ROC Curve analysis. Overview of the associations between MACE and the assessed clinical and immunological parameters within the HF cohort in a ROC curve analysis. CD4 and CD8 subset frequencies are always referred to their respective CD4 or CD8 parent population. * - HR is related to 1000 units of the MFI or the logarithmic NTproBNP levels; ** - Gives the number of patients where the investigators had information about the given parameter e.g. presence of absence of a disease/assist device; *** - Dichotomized Hazard ratio and p-values; NTproBNP – N-terminal prohormone of brain natriuretic peptide; TCM – T central memory cell; TEM – T effector memory cell; TEMRA – T effector memory CD45RA+ cell; Treg – T regulatory cell; Th – T helper cell; TC – T cytotoxic cell; MFI – Mean fluorescence intensity

| HFREF cohort MACE ROC analysis (N=113) |  |  |  |  |
| --- | --- | --- | --- | --- |
| Immune and clinical parameters | Patient count per Parameter** | Area under the curve | Standard Error | p-value |
| NTproBNP - Log * | 106 | 0.6645 | 0.0551 | 0.0051 |
| Hemoglobin - g/dl | 113 | 0.6674 | 0.0544 | 0.0100 |
| NTproBNP - pg/mL | 106 | 0.6645 | 0.0551 | 0.0051 |
| CD4 TEM Frequency >24% *** | 113 | 0.6530 | 0.0539 | 0.0054 |
| CD4 TEMRA Frequency - % | 113 | 0.6090 | 0.0548 | 0.0551 |
| CD4 TCM Frequency - % | 113 | 0.4002 | 0.0557 | 0.0788 |
| CD8 TCM Frequency - % | 113 | 0.3333 | 0.0970 | 0.1085 |
| Leukocyte - count/nl blood | 113 | 0.5790 | 0.0559 | 0.1641 |
| CD8 Effector/Memory Frequency - % | 113 | 0.6353 | 0.0999 | 0.1926 |
| CD8 Naive Frequency - % | 113 | 0.3667 | 0.1001 | 0.1992 |
| Thrombocyte - count/nl blood | 113 | 0.4323 | 0.0562 | 0.2330 |
| CD4 Naive Frequency - % | 113 | 0.4349 | 0.0563 | 0.2520 |
| CD4 Effector/Memory cell Frequency - % | 113 | 0.5651 | 0.0563 | 0.2520 |
| Urea - mg/dl | 107 | 0.5640 | 0.0583 | 0.2717 |
| Th17 Frequency - % | 113 | 0.5606 | 0.0572 | 0.2862 |
| estimated Glomerular Filtration Rate (MDRD) | 113 | 0.4401 | 0.0571 | 0.2919 |
| Th1 CD195 MFI* | 109 | 0.5588 | 0.0584 | 0.3108 |
| Creatinine Kinase Serum - U/l | 112 | 0.4423 | 0.0580 | 0.3126 |
| TroponinT - pg/ml | 94 | 0.5638 | 0.0720 | 0.3683 |
| CD8 TEM Frequency - % | 113 | 0.5922 | 0.1059 | 0.3749 |
| Low Density Lipoprotein - mg/dl | 113 | 0.5473 | 0.0573 | 0.4082 |
| CD8 TEM CD195 MFI* | 113 | 0.5440 | 0.0576 | 0.4416 |
| CD8 TEMRA Frequency - % | 113 | 0.5765 | 0.1062 | 0.4615 |
| Th22 Frequency - % | 113 | 0.5396 | 0.0578 | 0.4856 |
| Th1/17 CD195 MFI* | 110 | 0.5399 | 0.0588 | 0.4908 |
| Th9 Frequency - % | 113 | 0.5378 | 0.0564 | 0.5056 |
| Th1/17 Frequency - % | 113 | 0.5362 | 0.0565 | 0.5239 |
| Th9 CD195 MFI* | 110 | 0.5349 | 0.0590 | 0.5466 |
| High Density Lipoprotein - mg/dl | 113 | 0.5308 | 0.0563 | 0.5902 |
| CD4 TEMRA CD195 MFI* | 112 | 0.5278 | 0.0578 | 0.6248 |
| CD4 Naive CD195 MFI* | 112 | 0.4742 | 0.0575 | 0.6496 |
| CD4 TCM CD195 MFI* | 113 | 0.4763 | 0.0577 | 0.6770 |
| CD8 TEMRA CD195 MFI* | 113 | 0.5227 | 0.0578 | 0.6910 |
| Cholesterol - mg/dl | 113 | 0.5200 | 0.0582 | 0.7268 |
| Th2 Frequency - % | 113 | 0.4806 | 0.0590 | 0.7333 |
| Th22 CD195 MFI* | 110 | 0.5190 | 0.0591 | 0.7430 |
| Tc9 CD195 MFI* | 112 | 0.5183 | 0.0572 | 0.7485 |
| Th17 CD195 MFI* | 110 | 0.5157 | 0.0593 | 0.7859 |
| Left Ventricular End-Diastolic Diameter - mm | 111 | 0.4845 | 0.0592 | 0.7875 |
| CD8 Tc9 Frequency - % | 32 | 0.5275 | 0.1040 | 0.7915 |
| CD8 Naive CD195 MFI* | 113 | 0.4870 | 0.0567 | 0.8199 |
| CD4 TEM CD195 MFI* | 113 | 0.5116 | 0.0574 | 0.8375 |
| Th2 CD195 MFI* | 110 | 0.5106 | 0.0589 | 0.8554 |
| CD8 TCM CD195 MFI* | 113 | 0.4901 | 0.0572 | 0.8626 |
| Th1 Frequency - % | 113 | 0.4921 | 0.0579 | 0.8896 |
| Left Ventricular Ejection Fraction - % | 113 | 0.4959 | 0.0572 | 0.9430 |
| Treg Frequency - % | 113 | 0.5038 | 0.0588 | 0.9472 |

